# Diffusion Tensor Phenomapping of the Healthy and Pressure-Overloaded Human Heart

**DOI:** 10.1101/2024.05.03.24306781

**Authors:** Christopher A. Rock, Y. Iris Chen, Ruopeng Wang, Anne L. Philip, Boris Keil, Rory B. Weiner, Sammy Elmariah, Choukri Mekkaoui, Christopher T. Nguyen, David E. Sosnovik

## Abstract

Current techniques to image the microstructure of the heart with diffusion tensor MRI (DTI) are highly under-resolved. We present a technique to improve the spatial resolution of cardiac DTI by almost 10-fold and leverage this to measure local gradients in cardiomyocyte alignment or helix angle (HA). We further introduce a phenomapping approach based on voxel-wise hierarchical clustering of these gradients to identify distinct microstructural microenvironments in the heart. Initial development was performed in healthy volunteers (n=8). Thereader, subjects with severe but well-compensated aortic stenosis (AS, n=10) were compared to age-matched controls (CTL, n=10). Radial HA gradient was significantly reduced in AS (8.0±0.8°/mm vs. 10.2±1.8°/mm, p=0.001) but the other HA gradients did not change significantly. Four distinct microstructural clusters could be identified in both the CTL and AS subjects and did not differ significantly in their properties or distribution. Despite marked hypertrophy, our data suggest that the myocardium in well-compensated AS can maintain its microstructural coherence. The described phenomapping approach can be used to characterize microstructural plasticity and perturbation in any organ system and disease.

## INTRODUCTION

Diffusion tensor MRI (DTI) of the heart allows the microstructure of the myocardium, which varies markedly from endocardium to epicardium, to be measured noninvasively.^1–5^ However, the resolution of most translational DTI techniques provides only 4-5 voxels across the myocardium without interpolation,^6–10^ which likely under-resolves important features. Here, we present an approach to improve the spatial resolution of cardiac DTI by almost 10-fold. The key elements of the approach include the use of a cardiac-tailored 64-channel radiofrequency coil,^11^ a spatially-tailored 2D excitation pulse,^12^ a free-breathing sequence in which the diffusion-encoding gradients are designed to null the first two moments of motion,^13–15^ and respiratory gating using a low-rank tensor multitasking approach.^12, 16^

The submillimeter (sub-mm) resolution of the diffusion-encoded images was leveraged to measure local gradients in the cardiomyocyte helix angle (HA). At sub-mm resolution, HA in the led ventricle (LV) varied not only by radial depth across the myocardium but also by radial angle. Local gradients in HA were measured and used to compare the microstructure of the LV in patients with severe aortic stenosis (AS) and age-matched controls (CTL). In addition, a phenomapping matrix was developed that allowed voxel clusters with distinct microstructural properties to be identified. Principal component and cluster analysis of this matrix was performed to characterize the myocardium with an unheralded level of depth and determine whether microstructural differences existed between the CTL and AS subjects.

## RESULTS

### Cardiac DTI with Submillimeter Resolution

The development of sub-mm cardiac DTI was performed in a cohort of healthy volunteers (n=8). DTI scans were performed with a true (non-interpolated) in-plane resolution of 0.85mm x 0.85mm (Fig. 1a) followed immediately by an acquisition at the conventional (2.5mm x 2.5mm) resolution (Fig. 1b). The led anterior descending (LAD) coronary artery was routinely visible in single-average b50 and b500 images at 0.85mm resolution but became blurred at standard (2.5mm) resolution (Fig. 1a-b). To determine whether the improved resolution produced the expected gains in image sharpness, the LAD was used as a point source (supplement, Figure S1) for quantitative evaluation (n=7). Mean LAD diameter was significantly smaller at 0.85mm resolution for both the b50 (2.6±0.3mm vs. 5.0±1.5mm, p=0.003, paired t-test, Fig. 1c) and b500 (3.0±1.0mm vs. 5.6±1.4mm, p=0.003, paired t-test, Fig. 1d) images. Similarly, sharpness was significantly higher at 0.85mm resolution for both the b50 (0.55±0.08mm^-1^ vs. 0.29±0.08mm^-1^, p=0.003, Fig. 1e) and b500 (0.51±0.08mm^-1^ vs. 0.30±0.07mm^-1^, p<0.001, Fig. 1f) images.

**Figure 1.**
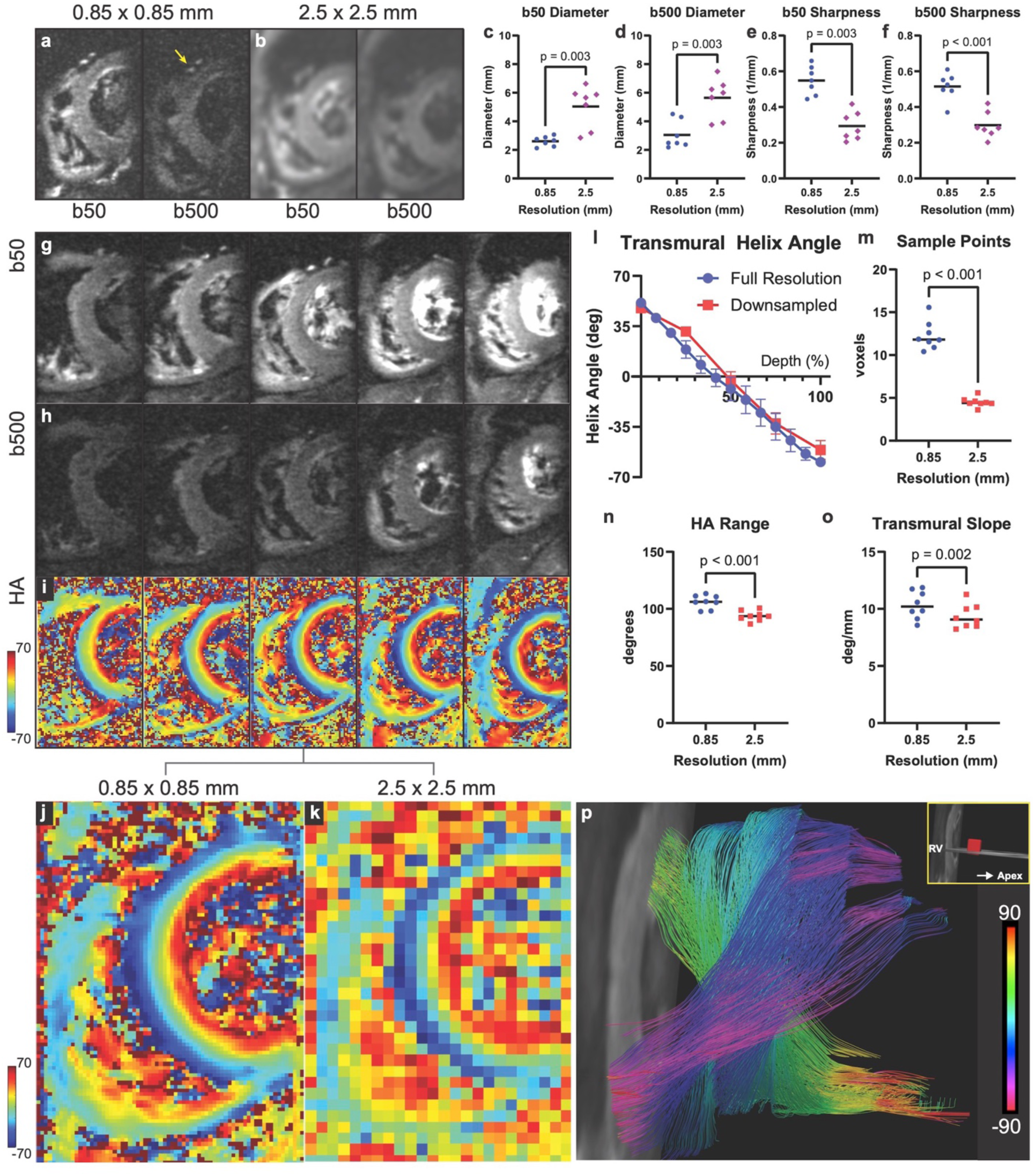
Diffusion tensor MRI (DTI) of the human heart with submillimeter resolution. Images were acquired with a 64-channel cardiac coil, a spatially-tailored 2D-excitation pulse and an in-plane resolution of 0.85 x 0.85mm. (a) Single-average short axis images of the heart acquired with low (b50) and high (b500) levels of diffusion encoding. The led anterior descending coronary artery (LAD, yellow arrow) can be clearly visualized and differentiated from the adjacent great cardiac vein. (b) Images acquired at standard resolution (2.5 x 2.5mm), in which the vessels appear blurred and cannot be clearly resolved. (c-f) Quantitative analysis using the LAD as a point source. Vessel diameter was accurately resolved at 0.85mm resolution in both the (c) b50 and (d) b500 images but significantly distorted at 2.5mm resolution (2.6±0.3mm vs. 5.0±1.5mm, p=0.003; 3.0±1.0mm vs. 5.6±1.4mm, p=0.003, paired t-tests, n=7). Likewise, vessel sharpness at both (e) b50 and (f) b500 was significantly higher in the 0.85mm images (0.55±0.08mm^-1^ vs. 0.29±0.08mm^-1^, p=0.003; 0.51±0.08mm^-1^ vs. 0.30±0.07mm^-1^, p=0.003, paired t-tests, n=7). (g-i) Free-breathing DTI of the entire heart in a healthy volunteer at 0.85 x 0.85 mm resolution. Single-average (g) b50 and (h) b500 images of 5 contiguous short axis slices are shown. (i) Corresponding helix angle (HA) maps of these slices (0.85 x 0.85mm, 12 averages). (j, k) Magnified view of middle HA map at (j) original resolution (0.85mm) and (k) downsampled to the standard resolution (2.5mm). The myocardium at the standard resolution appears highly pixelated and under-resolved. (l) Plot of HA in the septum vs. %-transmural depth at the high and standard resolutions. The standard resolution curve is less linear and has only 5 points across the myocardium. (m-n) Parameters of HA evaluated at 0.85mm vs. 2.5mm resolution in 8 healthy volunteers. (m) The number of transmural sample points in the HA plots averaged 12.3±1.6 at 0.85mm resolution vs. 4.5±0.6 at 2.5mm resolution (p<0.001, paired t-test, n=8). HA range across the septum was significantly higher at 0.85mm resolution (106±6° vs. 94±4°, p < 0.001, paired t-test, n=8). Likewise, the transmural slope (deg/mm) of HA was significantly higher at 0.85mm resolution (10.3±1.2°/mm vs. 9.4±1.0°/mm, p=0.002, paired t-test, n=8). (p) DTI tractography at 0.85 x 0.85mm resolution. Tracts passing through a small region-of-interest in the septum (red cube, inset) are visualized from the right ventricle. The crossing helical architecture of the subendocardial and subepicardial tracts, and circumferential orientation of the midmyocardial tracts, is well resolved.

Free-breathing DTI of the entire led ventricle (LV) was performed using 12 short axis slices. A stack of single average b50 (Fig. 1g) and b500 (Fig. 1h) images is shown in Fig. 1 and demonstrates the high quality of the images at 0.85mm resolution. Helix angle (HA) maps were generated using 12 averages (Fig. 1i) and consistently showed the expected evolution in HA from the endo to epicardium.^1–5^ To evaluate the impact of the improved resolution, 3-5 HA maps at the midventricular level were selected in each subject (n=8) and downsampled to the standard (2.5mm) resolution. In each subject, the values in the selected slices were combined to yield a single value for that subject. While the HA maps at 0.85mm resolution appeared sharp, those at 2.5mm resolution were noisy and highly pixelated (Fig. 1j-k). Plots of HA vs. % transmural depth were highly linear at 0.85mm resolution but not at 2.5mm resolution (Fig. 1l). The number of sampling points (voxels) from endo to epicardium in the HA plots averaged 12.3±1.6 at 0.85mm but only 4.5±0.6 at 2.5mm (Fig. 1m, p<0.001). The transmural range in HA was significantly higher at 0.85mm vs. 2.5mm resolution (106±6° vs. 94±4°, p=0.002, Fig. 1n), and the transmural HA slope was also significantly higher (10.3±1.2°/mm vs. 9.4±1.0°/mm, p<0.001, Fig. 1o). The 0.85mm resolution DTI datasets also successfully supported analysis with DTI tractography (Fig. 1p).

### Derivation of Local Helix Angle Gradients

We next imaged a group of subjects with severe aortic stenosis (AS, n=10) and a group of age-matched controls (CTL, n=10). We again acquired 12 short axis slices from base-apex but limited the analysis to 3-5 slices at the midventricular level (Fig. 2a). The rationale for this is demonstrated in the plot of single-average signal-to-noise (SNR) values from base to apex, shown in Fig. 2b. As proximity to the 64-channel surface coil increases, SNR grows producing the highest signal in the mid-LV and apex (see also supplement, Figure S2). Analysis of the most apical slices, however, can be complicated by through-plane motion during free-breathing.^12^ High quality HA maps could be consistently acquired at the midventricular level and the analysis was, thus, performed in these 3-5 slices. HA maps in a CTL and an AS subject are shown in Fig. 2c-d. The increased thickness of the myocardium in AS can be easily appreciated but the transmural pattern of HA appears, qualitatively, very similar to CTL.

**Figure 2.**
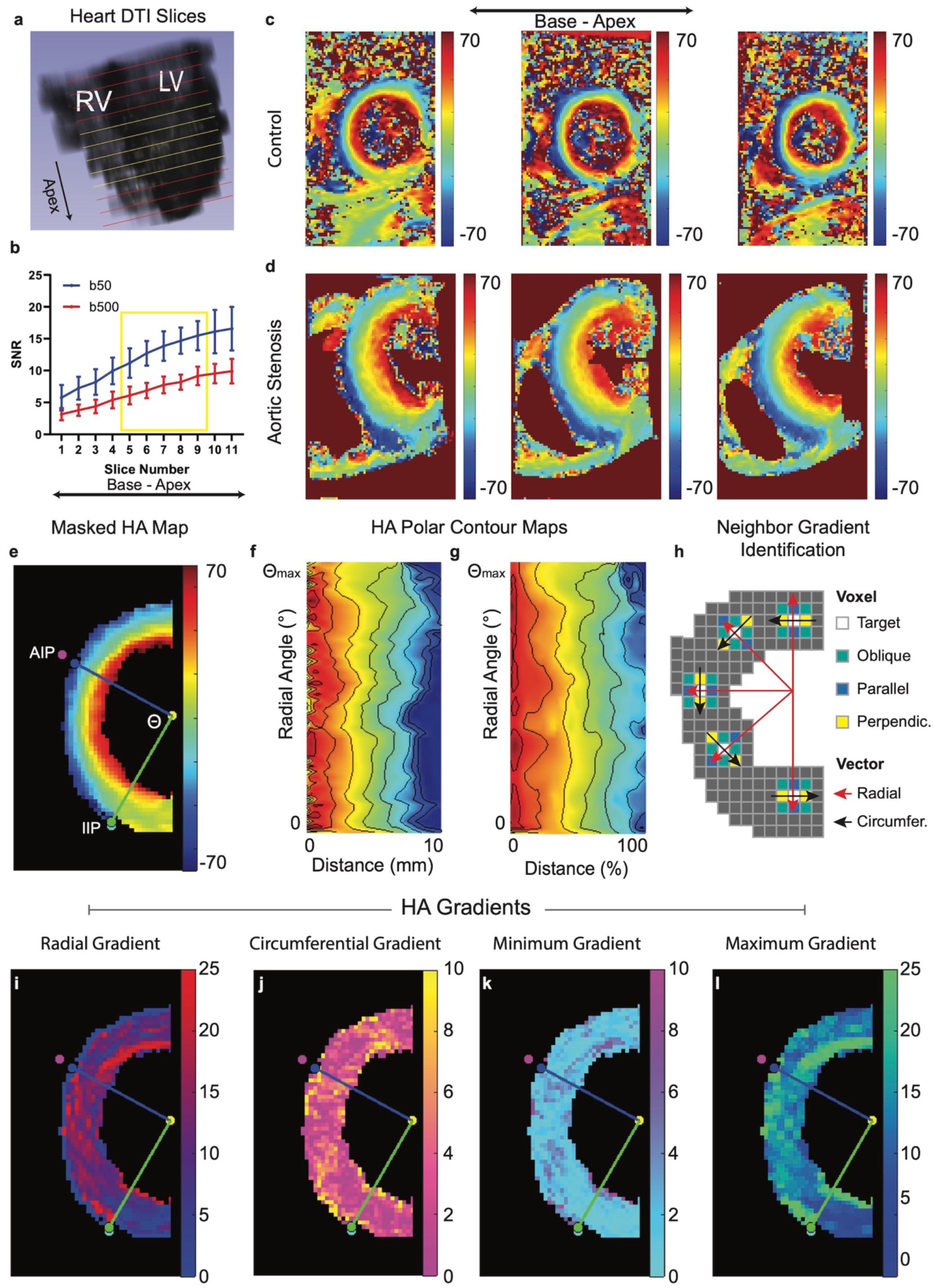
Measurement of local gradients in high resolution helix angle maps. (a). 3D reconstruction of the heart showing the location of the acquired DTI images. LV = led ventricle, RV = right ventricle. (b) Single-average signal-to-noise ratio (SNR) values (mean ± standard deviation) in the septum of healthy volunteers (n=6) from the most basal to apical slice. SNR was highest at the apex but overall image quality was highest in the 5-6 slices at the mid-ventricular level (yellow lines panel A). Analysis of the data was, therefore, performed in each subject using 3-5 midventricular slices. (c, d) High resolution HA maps of 3 midventricular slices in (c) an age-matched control (CTL) and (d) a subject with aortic stenosis (AS). The LV in AS is markedly thickened but qualitatively shows a similar HA pattern to CTL. (e) Masked HA map in CTL subject with the endocardial/epicardial trabeculations and RV removed. Radial sampling lines from the center of the LV to the anterior RV insertion point (AIP) and inferior RV insertion point (IIP) are shown. Analysis of the HA maps was performed using at least 22 radial sampling lines between the 2 insertion points. (f-g). Contour maps of HA, where data between the two insertion points in panel E are plotted as a function of radial distance (r) along a sample line and the radial angle (θ) of that line. The AIP defines a zero radial angle and radial distance can be defined in (f) mm or (g) as %-transmural depth. The HA contour maps reveal that substantial variations in HA exist both as a function of radial distance and radial angle, motivating the creation of a scheme to measure local HA gradients. (h) Definition of local HA gradients. Radial HA gradients occur between the target voxel and adjacent parallel voxels on a radial vector line (red). Circumferential HA gradients occur between the target voxel and adjacent perpendicular voxels on a circumferential vector line (black). (i-l) Local HA gradients in an age-matched control derived from the HA map shown in panel E. Maps of (i) radial HA gradient, (j) circumferential HA gradient, (k) minimum HA gradient and (l) maximum HA gradient are shown. All maps are at 0.85 x 0.85mm resolution.

Analysis of the HA maps was performed in the septum, where image quality was highest. The analysis region was demarcated by radial lines extending from the center of the LV to the anterior RV insertion point (AIP) and the inferior insertion point (IIP) across a radial angle 8, which is most cases approached ν/2 (Fig. 2e). The voxels in the septum were mapped using polar coordinates, where the position of each voxel is a function of its radial angle (8) from the anterior insertion point and its radial distance (r) from the endocardial boundary, expressed either as an absolute distance or as percent transmurality. A conventional HA map and polar contour HA maps in a CTL subject are shown in Fig. 2e-g. The magnified polar maps underscore that HA varies not only as a function of radial depth but also as a function of radial angle along circumferential contour lines. In any given voxel, radial and circumferential vectors can be defined (Fig. 2h) and local gradients in HA can be calculated along these unit vectors. Additional gradients can be calculated along oblique vectors to other neighboring voxels. In each voxel the radial (Fig. 2i), circumferential (Fig. 2j), minimum (Fig. 2k) and maximum (Fig. 2l) HA gradients could, therefore, be measured. Representative maps of these gradients, derived from the CTL HA map shown in panel E, are shown in Fig. 2i-l.

### Evaluation of LV Microstructure in Aortic Stenosis

#### HA Slope and Variance

LV microstructure in the AS (n=10) and CTL (n=10) subjects was initially compared by measuring HA slope and variance. Hypertrophy of the myocardium in AS results in a significant increase in the number of voxels across the septum and, hence, a reduction in the HA range per voxel. To determine whether this affected HA variance, the HA maps in the AS subjects were analyzed at their original resolution (0.85mm) and at a downsampled resolution (ASmatch) to match the HA range per voxel of the CTL group. The average downscaling factor used in the ASmatch analysis was 1.4x, which corresponds to a resolution of 1.2mm. HA maps of a CTL and AS subject are shown in Fig. 3a-b and the ASmatch HA map in Fig. 3c. Corresponding contour maps (Fig. d-i) and transmural HA profiles (Fig. 3j-o) are shown in the same row of each HA map. The contour maps and transmural plots show that the evolution of HA across the myocardium is highly linear, particularly when HA is plotted vs. %-transmural thickness.

**Figure 3.**
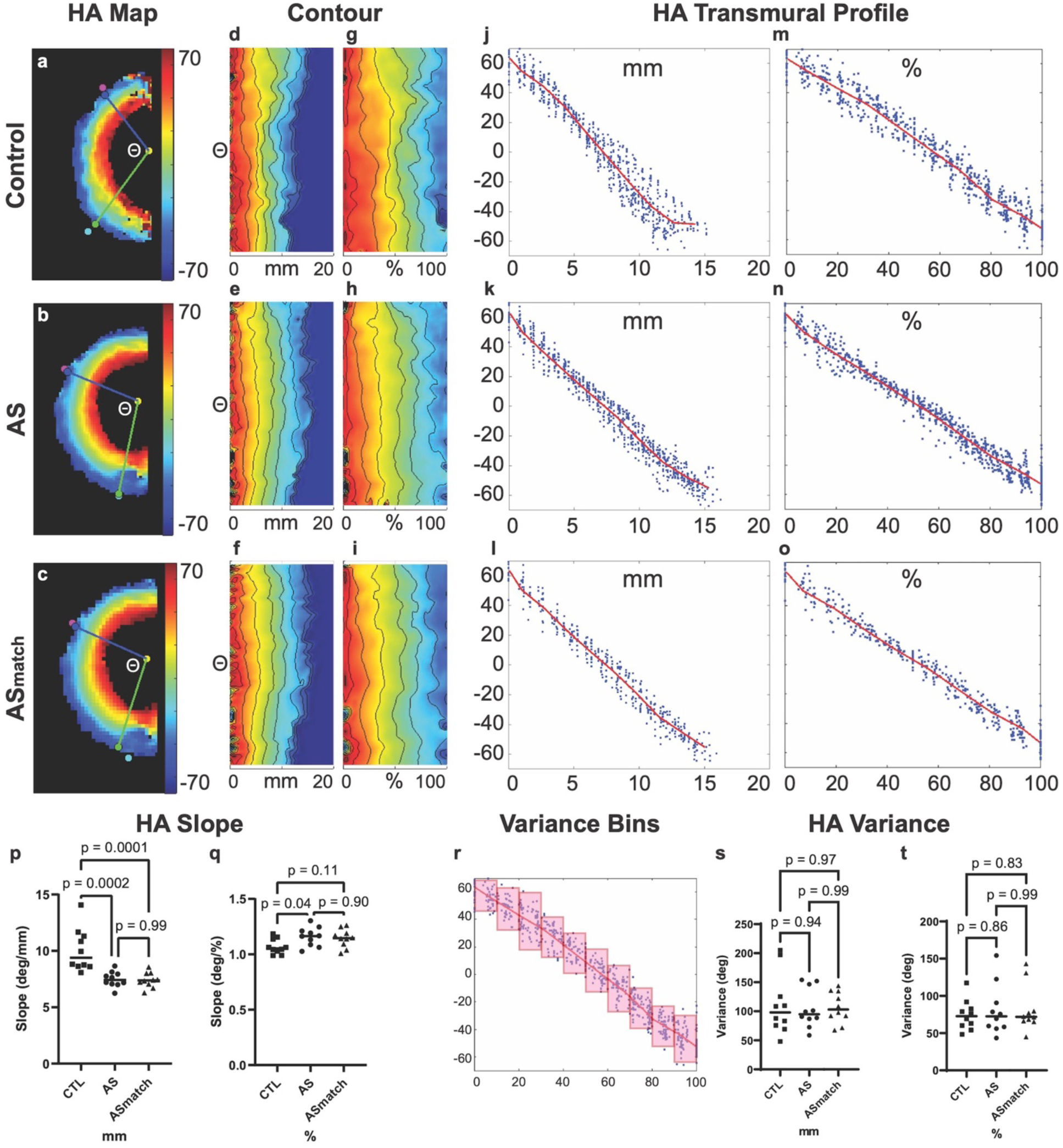
Impact of aortic stenosis on helix angle slope and variance. (a, b) HA maps in an age-matched control (CTL) and subject with severe aortic stenosis (AS). (c) HA map of the AS subject downsampled by a factor of 1.4 to match the number of voxels from endo to epicardium, and hence HA range/voxel, to that of the age-matched controls. (d-i) Contour maps of the septum show that the microstructure of the heart remains ordered in severe AS despite a marked increase in myocardial thickness. The contour lines show similar variations in the CTL and AS maps, both when radial position is classified by distance (d-f) or by %-transmurality (g-i). In addition, no visible differences were produced by downsampling to produce the ASmatch maps. (j-o). Transmural profiles of HA in the maps shown in panels A-C. HA values in individual voxels are plotted vs. (j-l) absolute or (m-o) %-transmural depth. The transmural slope of HA is highly linear, particularly when plotted as a function of %-transmural depth. (p) HA slope was reduced in the AS subjects when calculated per unit distance in mm (7.5±0.7deg/mm vs. 10.1±1.9deg/mm, p=0.002, Anova, n=10 per group). (q) However, when calculated vs. %-transmural depth, HA slope was significantly increased in the AS subjects (1.16±0.08deg/% vs. 1.07±0.07deg/%, p=0.044, Anova, n=10 per group). (p, q) No differences in HA slope were seen between the AS and ASmatch datasets. (r) Schematic of HA variance calculation. Voxels were binned to create 10 populations per profile, and HA variance in each bin was calculated and then averaged. (s, t) No differences were seen in HA variance between the CTL, AS and ASmatch cohorts (n=10 per group, Anova) both when plots of HA vs. depth in mm (s) and plots of HA vs. %-transmural depth (t) were used.

HA slope per mm was significantly reduced in AS vs. CTL (7.5±0.7deg/mm vs. 10.1±1.9deg/mm, p=0.002, Fig. 3p). Conversely, HA slope per %-transmurality was significantly increased in AS vs. CTL (1.16±0.08deg/% vs. 1.07±0.07deg/%, p=0.044, Fig. 3q). No significant differences (p>0.9) were seen between the AS and ASmatch HA maps. HA variance was calculated by dividing each HA curve into 10 bins organized by transmural depth (either in mm or %-transmurality), calculating the variance in each bin (Fig. 3r), and then averaging the values in all slices to produce a single HA variance value per subject. No significant differences (p>0.8) were seen in HA variance between the AS and CTL subjects (Fig. 3s-t). Likewise, no differences (p>0.8) in HA variance were seen between the AS and ASmatch maps. Collectively, these data show that the profound hypertrophy of the LV in AS is not accompanied by a loss of microstructural coherence.

#### Local HA Gradients

To evaluate LV microstructure in more detail, we next compared local HA gradients, in deg/mm, between the AS (n=10) vs. CTL (n=10) subjects. The echo time used to acquire the DTI data was 76ms and, consequently, the b50 images provided a T2-weighted signal map. Maps of b50, mean diffusivity (MD), fractional anisotropy (FA), HA and radial HA gradient are shown in a CTL (Fig. 4a-e) and AS subject (Fig. 4f-j). The myocardium in the b50 images, which were acquired in midsystole, was significantly thicker in AS (15.4±1.4mm vs. 10.9±1.6mm, p<0.0001, Fig. 4k). No differences in MD (1.41±0.11*10^-3^mm^2^/s vs. 1.44±0.07*10^-3^mm^2^/s, p=0.72, Fig. 4l) or FA (0.335±0.021 vs. 0.325±0.017, p=0.49, Fig. 4m) were seen. HA range was not significantly increased in AS (110±8° vs. 106±6, p=0.56, Fig. 4n), however, radial HA gradient was significantly lower in AS vs. CTL (8.0±0.8°/mm vs. 10.2±1.8°/mm, p=0.001, Fig. 4o). No significant difference was seen in the circumferential gradient (3.78±1.1°/mm vs. 3.31±0.53°/mm, p=0.46, Fig. 4z) between AS and CTL. The impact of spatial resolution on scalar metrics of diffusivity and local HA gradients is shown in the supplement. No significant differences were seen between the AS and ASmatch datasets (supplement Figure S3), but significant differences in all metrics were seen between 0.85mm vs. 2.5mm resolution (supplement, Figure S4).

**Figure 4.**
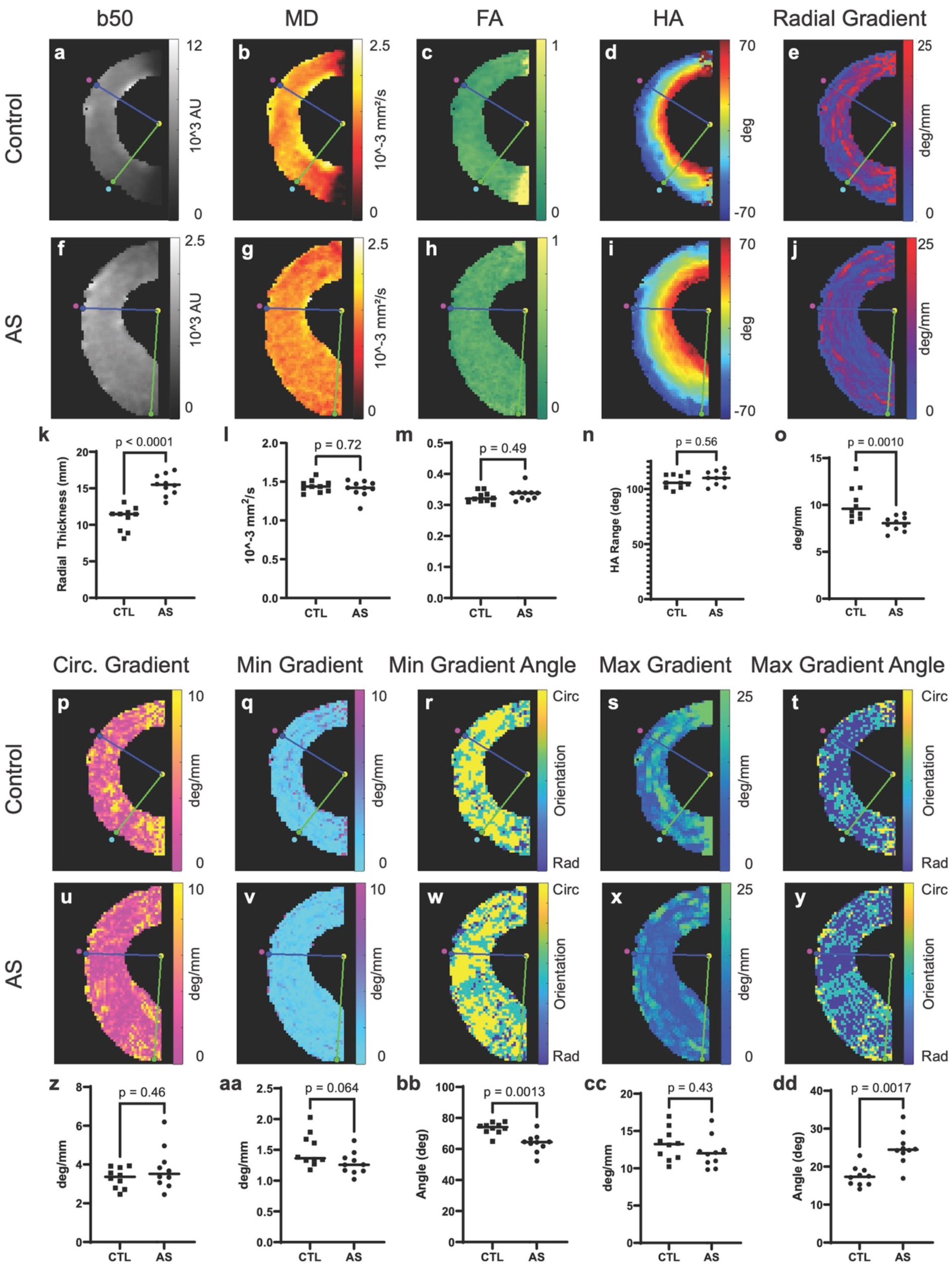
Local HA gradients in patients with aortic stenosis. (a-j) b50 images, mean diffusivity (MD), fractional anisotropy (FA), HA and radial HA gradient maps in an aged-matched CTL (n=10) and subject with AS (n=10) are shown. (k) Myocardial thickness in the b50 images, acquired in midsystole, was significantly increased in the AS subjects (15.4±1.4mm vs. 10.9±1.6mm, t-test, p=<0.001). No differences were seen in MD (l) and FA (m) between the AS and CTL subjects. (n) HA range trended upwards in the AS subjects but did not reach significance. (o) A significant decrease (8.0±0.8°/mm vs. 10.2±1.8°/mm, p=<0.001) in radial HA gradient was seen in the AS subjects. (p-y) Additional gradient maps from the same CTL and AS subjects are shown including circumferential HA gradient, minimum HA gradient, minimum gradient angle, maximium HA gradient and maximum gradient angle. (z-aa) There was no significant difference in circumferential HA gradient (3.78±1.1°/mm vs. 3.31±0.53°/mm, p=0.46), but a strong trend towards a lower minimum HA gradient was seen in AS (1.28±0.18°/mm vs. 1.50±0.27°/mm, p=0.064). (bb) The angle between the voxels forming the minimum HA gradient was significantly less circumferential in AS (64±6° vs. 73±4°, p=0.0013). (cc) No significant differences were seen in maximum HA gradient between AS vs. CTL (12.1±2.1°/mm vs. 13.2±2.1°/mm, p=0.43). (dd) However, the direction of the maximum HA gradient was significantly less radial in AS (24.8±4.3° vs. 17.4±2.6°, p=0.0017). Gradients in the CTL, AS and ASmatch groups were compared with Anova and Tukey’s post-test comparison. No significant differences were present between the AS and ASmatch groups (see supplement).

The orientation of the minimum (min) and maximum (max) HA gradients, relative to the radial vector, was also recorded. If the min/max gradient subtended an angle of 0-30° this was regarded as radial, 30-60° as oblique, and 60-90° as circumferential. The min HA gradient trended strongly to a reduced value in AS (1.28±0.18°/mm vs. 1.50±0.27°/mm vs, p=0.064, Fig. 4aa), but there were no significant differences in the max HA gradient (12.1±2.1°/mm vs. 13.2±2.1°/mm vs, p=0.43, Fig. 4cc). The min gradient was less circumferential in AS (64±6° vs. 73±4°, p=0.0013, Fig. 4bb) and the max gradient was less radial (24.8±4.3° vs. 17.4±2.6°, p=0.0017, Fig. 4dd). These differences were largely a reflection of the lower radial HA gradient in AS (supplement, Figure S5). Collectively, the HA gradient maps confirmed that the hypertrophied myocardium in well compensated AS maintains, and may even increase, its microstructural coherence.

### Microstructural and Pathophysiological Correlations in AS

We next sought to determine the correlations, on a per subject basis, between metrics of LV microstructure and commonly used clinical variables (Fig. 5a). The metrics with the highest and most consistent correlations included diastolic LV thickness, HA variance, min gradient, and circumferential gradient. A strong correlation was seen between the min and circumferential HA gradients (Fig. 5b), particularly in AS. In the AS subjects an increase in diastolic LV thickness correlated with reduced HA variance (Fig. 5c), and reduced HA gradients (Fig. 5d-g). In addition, the correlations between HA variance and the local HA gradients were stronger in the AS than CTL subjects (Fig. 5h-k). This again suggests that that the hypertrophied LV in AS does not lose, and may actually increase, its microstructural coherence. DTI-tractography was performed in subjects with AS. A loss of tract coherence was seen in subjects with high HA variance and HA gradients (Fig. 5l-m). The clinical characteristics of the AS and CTL subjects are provided in Table 1 of the supplement.

**Figure 5.**
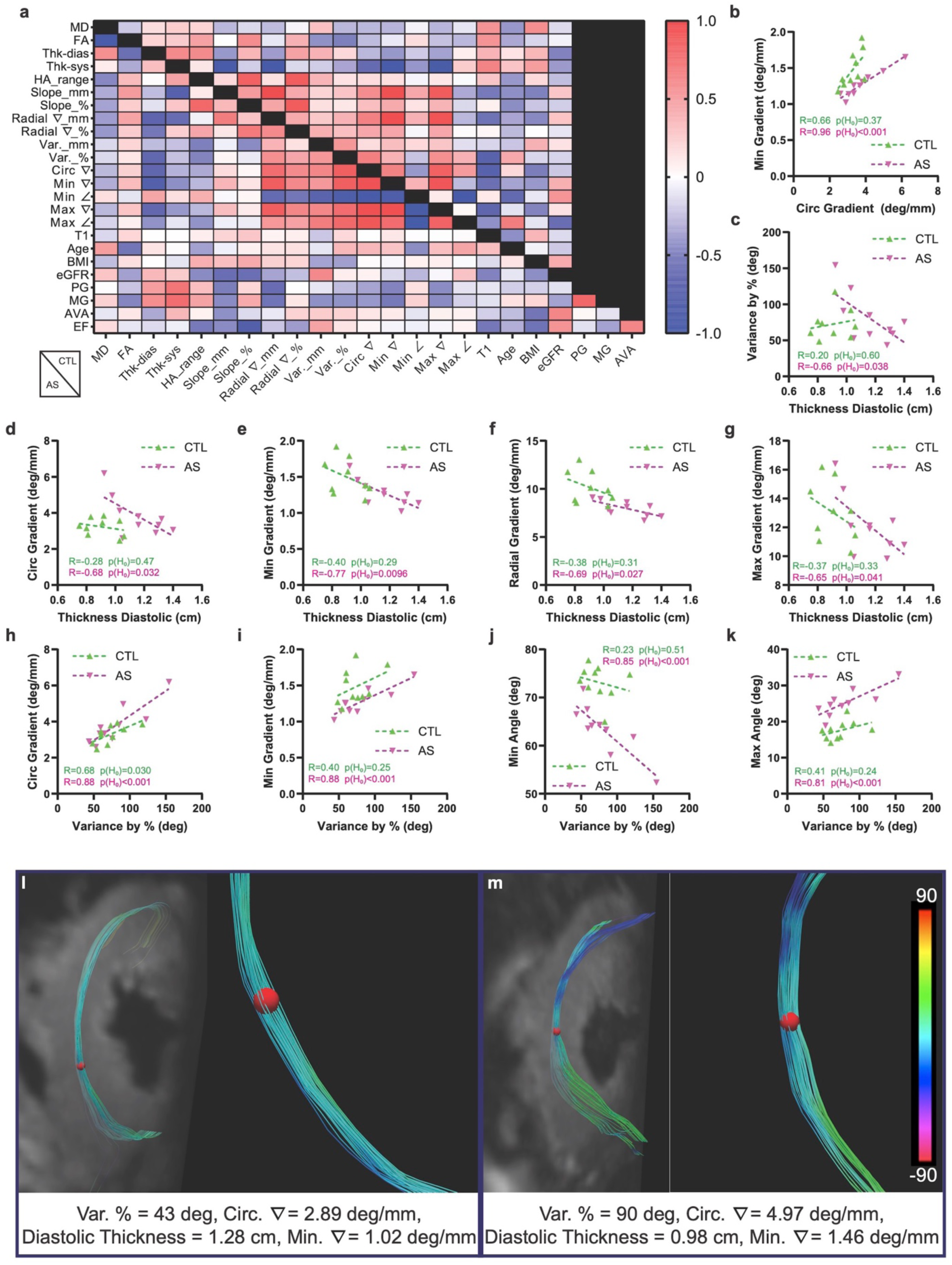
Correlation between measures of LV remodeling and microstructure. (a) Heatmap showing linear correlations (Pearson’s coefficient) between parameters of LV microstructure and remodeling in the CTL (above diagonal, n=10) and AS (below diagonal, n=10) subjects. In the AS subjects measures of LV load were included as well. (b) A strong correlation was seen between the minimum and circumferential HA gradients, particularly in the subjects with AS (R=0.96, p<0.001). (c-g) No significant correlations were seen between diastolic LV thickness (Thk-dias) and HA variance or any of the HA gradients in the CTL group. However, in the AS subjects, significant negative correlations were seen between diastolic LV thickness and (c) HA variance (p=0.038), (d) circumferential HA gradient (p=0.032), (e) minimum HA gradient (p=0.0096), (f) radial HA gradient (p=0.027), and (g) maximum HA gradient (p=0.041). (h) Significant correlations between HA variance and circumferential gradient were seen in both the CTL (p=0.030) and AS subjects (p<0.001). In the AS subjects HA variance also correlated with (i) minimum gradient (p<0.001), (j) minimum gradient angle (p < 0.001) and (k) maximum gradient angle (p<0.001). (l-m) Tractography in subjects with AS revealed that high HA gradients were associated with reduced tract coherence. Circumferential tracts intersecting a region-of-interest (red sphere) in the mid-myocardium are shown with magnified views of the tracts on the right. (l) AS subject with low HA variance and low HA gradients. The tracts in the midmyocardium are highly uniform and coherent. (m) AS subject with high HA variance and high HA gradients. The tracts in the midmyocardium show some loss of order and coherence. *Abbrevia(ons used in heatmap and figure are: MD (mean diffusivity), FA (frac(onal anisotropy), Thk-diast (diastolic LV thickness), Thk-sys (systolic LV thickness), HA_range (helix angle (HA) range), slope_mm (HA slope per mm), slope _% (HA slope per percent transmural thickness),* ∇*_mm = gradient/mm,* ∇*_% = gradient/% depth, Var. = variance,* ∇ *= gradient,* ∠ *= angle, T1 (longitudinal magne(c relaxa(on (me), BMI (body mass index), eGFR (es(mated glomerular filtra(on rate), PG (peak gradient across aor(c valve), MG (mean gradient across aor(c valve), AVA (aor(c valve area), EF (ejec(on frac(on)*.

### Diffusion Tensor Phenomapping

We next sought to evaluate the associations between metrics of LV microstructure on a per-voxel basis and determine whether distinct microstructural microenvironments were present. This approach is conceptually analogous to single cell RNA sequencing (supplement, Figure S6),^17, 18^ and has also been used to analyze complex clinical populations.^19^ In each subject, the voxels from all slices were collected and arranged into a subject data matrix (Fig. 6 a-c). Each voxel (row) in the matrix contained 10 values (columns) derived predominantly from the HA gradient maps (Fig. 6b). The values for each parameter (column) were normalized using a z-score to create a normalized subject data matrix. Prior to normalization the HA gradient values were square-root transformed to create highly Gaussian distributions (supplement, Figure S7). The normalized matrices of each CTL (n=10) and AS (n=10) subject were then combined to form a population data matrix (Fig. 6d), containing 33,458 rows (voxels). Principal component analysis (PCA) of the matrix showed that the first three principal components (PCs) accounted for >75% of the observed variation. The first axis (43% of variance) was defined primarily by high radial and max HA gradients, the second axis (19% of variance) by high MD and low FA, and the third axis (13% of variance) by high circumferential and min HA gradients (Fig. 6e).

**Figure 6.**
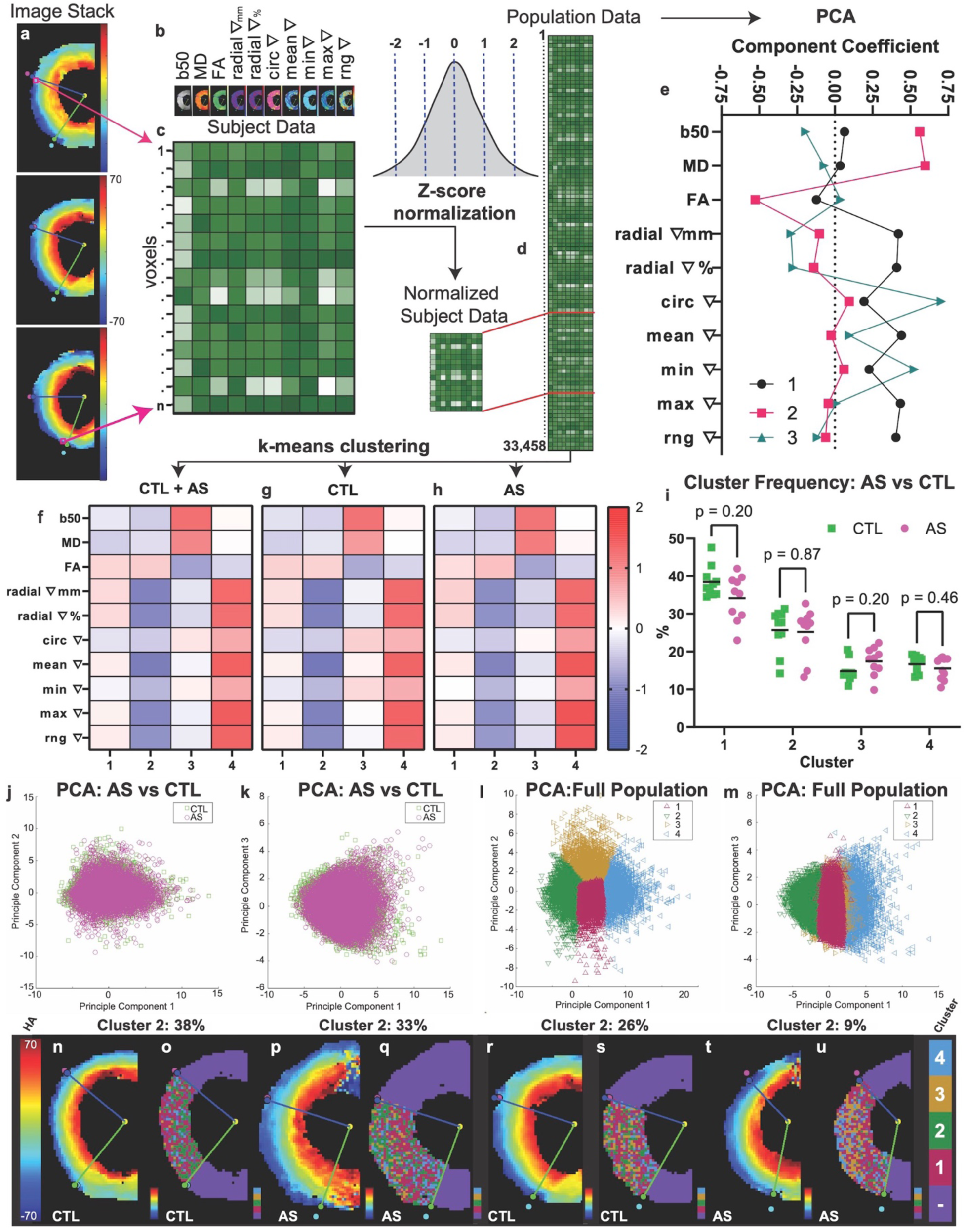
Diffusion tensor phenomapping of myocardial microstructure. The microstructural properties of each individual voxel in a stack of HA maps (a) were described by (b) a 10-element array. rngς = gradient range (maxς - minς). The arrays of all available voxels (1…n) in the stack were combined into (c) a single subject data matrix. Each parameter (column) in the matrix was then mapped onto a common scale via z-score normalization to yield a normalized subject data matrix. (d) The normalized matrices of all subjects (CTL=10, AS=10) were then combined to create a population data matrix (d) with > 30,000 voxels (rows). (e) Principal component analysis (PCA) of this matrix revealed that >75% of the variation lay within the first 3 principal components. (e) Principal component 1 (PC1) was dominated by the radial and maximum HA gradients, PC2 by scalar measures of diffusivity (MD, FA), and PC3 by circumferential and minimum HA gradients. (f) K-means clustering of the population matrix into 4 clusters. Cluster 1 consisted of voxels with average HA gradients and above average FA, cluster 2 of voxels with the lowest HA gradients and above average FA, cluster 3 of voxels with below-average HA gradients but above average MD and below average FA, and cluster 4 of voxels with below average FA and the highest gradients. (g, h) Dividing the full matrix into separate CTL and AS matrices produced cluster sets with extremely similar properties to each other (CTL vs. AS) and the full set (CTL+AS). (i) The percentage of voxels falling into each cluster was also similar in AS and CTL. PCA plots did not reveal any separation between voxels from AS (magenta circles) and CTL (green squares) subjects using (j) PC1 and PC2, or (k) PC1 and PC3. (l, m) PCA of voxels, classified by cluster, showed some separation of voxels in clusters 2 and 4 along PC1. (n-u) Cluster maps of voxels in the septum, depicting the microstructural cluster each voxel falls into, and their corresponding HA maps. (n-u) CTL and AS subjects with varying proportions of voxels in cluster 2 are shown. The HA maps in all 4 hearts appear similar but the proportion of highly ordered voxels (cluster 2) in them ranges from 38% to 9%.

k-means clustering was performed on the population matrix as a whole, and then separately on the voxels from the CTL or AS subjects only. A minimum of 4 voxel clusters were consistently identified, and the properties of corresponding clusters were extremely similar across all cases (Fig. 6f-h). The first cluster consisted of ordered voxels with average HA gradient values and FA values slightly above average. The second cluster consisted of highly ordered voxels with above average FA values and below average HA gradients. The third cluster contained voxels with average HA gradients but above average MD and below average FA. The fourth cluster contained the least ordered voxels with the highest HA gradients and below average FA. The proportion of voxels in each cluster did not differ between AS and CTL (Fig. 6i) and was 34±6% vs. 38±4% for cluster 1 (p=0.20), 25±6% vs. 26±6% for cluster 2 (p=0.87), 17±4% vs. 15±3% for cluster 3 (p=0.20), and 16±3% vs. 17±2% for cluster 4 (p=0.46). PCA plots showed no difference in the distribution of voxels from the AS and CTL populations (Fig. 6j-k) but could resolve voxels in clusters 2 from those in cluster 4 (Fig. 6l-m). Cluster maps were created by classifying each voxel in the septum by the cluster it belonged to. Cluster maps in CTL and AS subjects are shown with their corresponding HA maps in Figure 6n-u.

We next assessed the spatial distribution of the voxels in the 4 clusters, with particular focus on %-transmural depth. The total number of voxels (all clusters combined) increased linearly with %-transmural depth due an increase in circumference towards the epicardium (Fig. 7a-c). However, distinct transmural distributions were seen for all 4 clusters. The voxels in cluster 1 mirrored the transmural increase in voxel number but also showed a distinct prominence in the subepicardium (Fig. 7d-f). Those in cluster 2 had a bell-shaped distribution and were most concentrated in the midmyocardium (Fig. 7g-i). The voxels in cluster 3 showed a preference for the subendocardium and subepicardium (Fig. 7j-l) and those in cluster 4 for the subepicardium (Fig. 7m-o).

**Figure 7.**
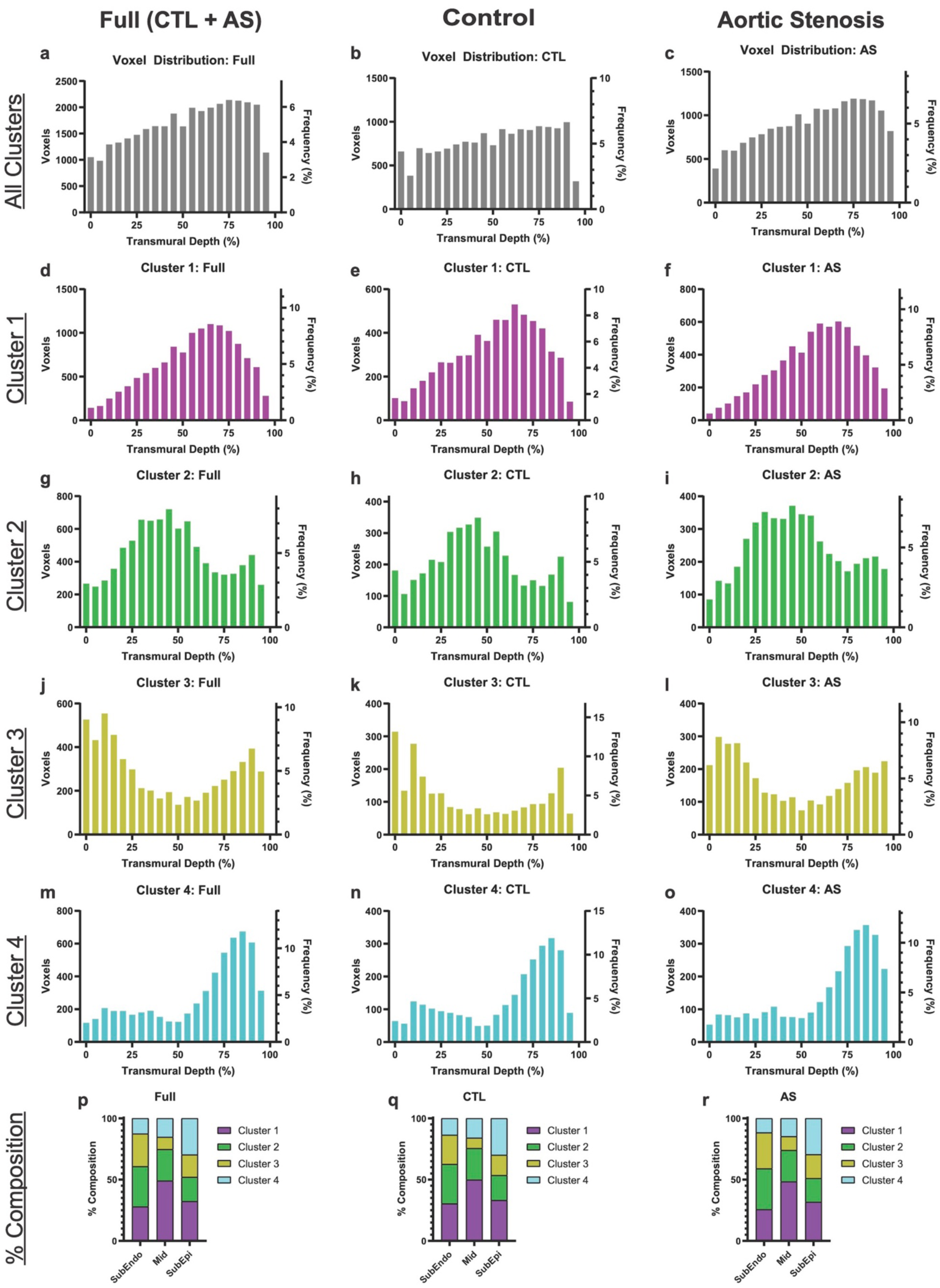
Spatial distribution of microstructural clusters. Histograms of voxel distribution as a function of transmural depth are shown for all clusters combined and each cluster individually. (a-c) The total number of voxels (all clusters combined) increased linearly from endocardium to epicardium due to a progressive increase in the circumference of the LV. (d-f) The voxels in cluster 1 (average order) showed the transmural increase seen with all the clusters combined, but also a slightly higher concentration in the subepicardium. (g-i) The distribution of voxels in cluster 2 (highly ordered) showed a bell-shaped distribution centered in the midmyocardium. (j-l) The voxels in cluster 3 were most concentrated in the subendocardium and subepicardium, and lowest in the midmyocardium. (m-o) Voxels in cluster 4 (least ordered) were distributed throughout the myocardium but showed a distinct peak in the subepicardium. (p-r) Proportion of voxel clusters across the layers of the heart. In the subendocardium there is a high proportion of voxels from cluster 2 (33%) and similar proportions of voxels from clusters 1 (28%) and 3 (27%). In the midmyocardium there is a high proportion of voxels from cluster 1 (49%) followed by cluster 2 (26%). In the subepicardium the proportion of voxels from cluster 2 decreased (20%), while that from cluster 4 increased (30%). No significant differences in the transmural distribution of the clusters (d-o) or their prevalence in the layers of the myocardium (p-r) were seen between the AS and CTL subjects.

The relative contribution of voxels from each cluster across the layers of the myocardium is shown in Figure 7p-r. In the subendocardium voxels in cluster 2 were the most prominent (33%), followed by similar proportions from cluster 1 (28%) and cluster 3 (27%). In the midmyocardium cluster 1 (49%) was the most dominant followed by cluster 2 (26%). In the subepicardium the proportion of voxels from clusters 1 (32%) and 2 (20%) decreased, while that from cluster 4 increased (30%). No significant differences in the overall breakdown of the clusters, their transmural distributions or their relative contributions in the layers of the myocardium were seen between the AS and CTL subjects. Collectively, PCA and cluster mapping did not reveal any loss of microstructural coherence in the AS subjects.

Additional details on the phenomapping approach are provided in the supplement. Specifically, the impact of subject vs. population-based normalization and k=4 vs. k=5 clustering is shown in supplemental Figure S8. Likewise, the impact of spatial resolution on the ability to perform voxel-based phenomapping is shown in supplemental Figure S9.

## DISCUSSION

We show in this study that free-breathing DTI of the human heart can be performed with sub-mm in-plane resolution. We further show that this enhanced resolution allows local gradients in cardiomyocyte orientation to be measured and used in a phenomapping approach to characterize local variations in the structural microenvironment of the heart. The approach was successfully implemented in subjects with severe AS, whose hearts are subjected to a major increase in load. However, despite marked hypertrophy of the myocardium, the microstructure of the heart in AS remained highly coherent and organized.

DTI of the heart can be performed with a diffusion-encoded spin echo approach, provided gradient waveforms designed to null the first two moments of motion are used.^13–15^ While an in-plane resolution as low as 1.6×1.6mm has been reported,^20^ we and most others have used an in-plane resolution of 2.3-2.5mm.^7, 12, 21–23^ A similar resolution (2.5-2.8mm in-plane) has been used in most DTI studies of the heart using a stimulated echo approach.^6, 8, 24–27^ The ability of diffusion-encoded spin echo images to be acquired during free breathing is of major appeal,^9, 12, 28^ however the images are vulnerable to severe distortion and susceptibility artifacts in the lateral wall.^28, 29^ Here, we leveraged the advantages of the spin echo approach and eliminated the disadvantages by selectively imaging those areas of the heart least affected by these artifacts. Combined with a high k-space matrix and a cardiac-tailored 64-element radiofrequency coil, focused DTI of most of the heart could be performed with a non-interpolated in-plane resolution of 0.85×0.85mm

The improved resolution of our DTI acquisition was leveraged to introduce a voxel-based phenomapping approach which, analogous to single cell RNA sequencing,^17, 18^ allows clusters of cells and distinct structural microenvironments in the myocardium to be detected. The same number of microstructural clusters could be identified in both the CTL and AS subjects. The voxels in the most ordered cluster (cluster 2) fell predominantly in the midmyocardium, where FA has previously been reported to be highest.^8^ The least ordered voxels (cluster 4) constituted 17% of all voxels and could represent microenvironments with focal areas or fibrosis, amyloid or other factors promoting cardiomyocyte misalignment. Preclinical studies will be needed to correlate phenomapping clusters with detailed histological examination of the myocardium.

The LV myocardium in the AS subjects underwent significant hypertrophy but maintained its microstructural integrity. A prior report of DTI in subjects with AS (at conventional spatial resolution) showed that the LV adapted to the increased load by increasing the transmural slope of HA.^22^ A similar increase in HA slope was observed in this study but no metrics of microstructural disorganization were increased in the AS subjects and no differences were seen in the prevalence and distribution of the voxel clusters. Several metrics of LV microstructure even showed improved coherence in AS. It should be noted that the subjects in our study had severe but well-compensated AS. All subjects had LV ejection fractions >57% and maximum velocities across the aortic valve <5ms^-1^. In the absence of symptoms current guidelines do not recommend aortic valve replacement in such subjects,^30, 31^ and the results of our study support this recommendation.

The approach we describe provides a paradigm shid in both DTI acquisition and analysis, but several limitations of the approach merit discussion. The focused FOV approach we use may not be suited to conditions that selectively affect the lateral wall and not the septum. However, in conditions that affect the entire LV, strong precedent exists for limiting quantitative analysis to the septum, and this is routinely done in MR-spectroscopy,^32^ and T1/T2* mapping of the myocardium.^33^ Preclinical studies in pressure-overloaded rodent hearts have shown either small or no regional variations in HA,^34, 35^ but this will need to be further investigated in humans. Ultimately, improved distortion correction techniques will need to be developed to allow the lateral wall to be imaged with equivalent accuracy to the septum. The acquisition time of the high-resolution datasets remains long. However, we have shown that an artificial intelligence (AI) based approach can be used to denoise DTI datasets,^23^ and this could potentially reduce the time required for the acquisition of high-resolution data. The size of our study was designed for technical development and first-in-human application. However, all metrics that trended towards significance suggested that microstructural coherence was maintained or increased in AS, further validating the conclusions of the study. We limited our analysis in this study to the primary eigenvector of the diffusion tensor, which reflects cardiomyocyte orientation.^36^ Future studies will be required to determine whether the secondary eigenvector, which reflects the sheetlet angle in the heart,^6, 37, 38^ can be accurately measured at sub-mm resolution and incorporated into the described phenomapping approach.

In summary, we present a technique to perform DTI of the human heart with sub-mm resolution and leverage this to measure local gradients in cardiomyocyte helix angle. In addition, we describe a phenomapping approach to identify voxel clusters with distinct microstructural properties. We show, using these techniques, that the myocardium in well-compensated AS maintains, and may even increase, its microstructural integrity. The phenomapping approach we describe characterizes the microstructure of the myocardium with an unheralded level of depth and provides important insights into the plasticity of the heart.

## METHODS

Three cohorts of subjects were studied: 1) A group of healthy volunteers (n=8), who were used for technical development of the high-resolution DTI approach, 2) a group of subjects with severe aortic stenosis (n=10), and 3) a group of age and gender matched healthy controls (n=10). The study was approved by the institutional review board of our institution (IRB2016P001898) and written consent was obtained from all participants in the study.

### Image Acquisition

All subjects were imaged on a commercial 3T MRI system, equipped with 80mT/m whole body gradients (Prisma, Siemens Medical). Imaging was performed using a cardiac-tailored 64-element array,^11^ and/or commercial 32-element array. A free-breathing diffusion-encoded spin echo echoplanar imaging (EPI) sequence, with diffusion-encoding gradients designed to null the first and second moments of motion was used (supplement, Figure S1). We have previously used this sequence to image the entire LV with an in-plane resolution of 2.5×2.5mm.^12^ Here the spatially-tailored 2D excitation pulse in the sequence was used to selectively excite an even smaller field-of-view (FOV) centered on the interventricular septum. Specific parameters included: FOV 23.7cm, phase FOV 24.5%, acquisition matrix 286×70, slice thickness 8mm, resolution 0.85×0.85mm, chemical fatsat, spin echo EPI readout, echo-train length 46, 12 slices interleaved, repetition Jme (TR) = 12RR, echo time (TE) =76ms, bandwidth 1165 Hz/pixel, b = 50 s/mm^2^ (2 averages), b = 500 s/mm^2^ (12 averages), 12 diffusion encoding directions. Retrospective respiratory gating of the diffusion-encoded images was performed using a low-rank multitasking approach, as previously described.

Additional images acquired in the cohort of healthy volunteers of included:

- Noise images of the heart at 0.85×0.85mm in-plane resolution using the 64-element cardiac array. This allowed SNR and CNR maps of the DTI sequence at 0.85×0.85mm resolution to be created.
- DTI of the heart and noise images at 0.85×0.85mm in-plane resolution using a commercial 32-channel receive coil. These were used to compare the SNR and CNR of the 64 and 32-element arrays.
- DTI of the heart at 2.5×2.5mm in-plane resolution using the 64-element cardiac array. This was used to compare the point-spread of the coronary artery at 2.5×2.5mm vs. 0.85x 0.85mm resolution.

Parameters for these sequences were as described above, but in the noise images the flip angle was set to zero and the TR to 2.6s. DTI at 2.5×2.5mm resolution was performed with 6 averages since the SNR in these larger voxels would be expected to be almost 10-fold higher. The FOV and image matrix (112×30) were adjusted to produce an in-plane resolution of 2.5×2.5mm, which resulted in a TE of 72ms. In selected healthy volunteers, breath-hold balanced steady state free precession (bSSFP) cine images of the led anterior descending coronary artery were acquired with an in-plane resolution chosen to match that of the 0.85×0.85mm DTI images. These cines were acquired with the 64-channel coil and the following parameters: FOV 320×268mm, slice thickness 8mm, flip angle 44°, matrix 384 x 322, resolution 0.83×0.83mm, TR = 40ms, TE = 1.44ms, 25 phases per RR, averages 1.

Additional images acquired in the aortic stenosis (AS) and age-matched control (CTL) subjects included:

- T1 maps of the heart using a modified Look-Locker imaging (MOLLI) sequence. Parameters included: FOV 360×306mm, slice thickness 8mm, flip angle 35°, matrix 256×218, resolution 1.4×1.4mm, TE = 1.12ms.
- bSSFP cines of the heart. Parameters included: FOV 339×284mm, slice thickness 6mm, flip angle 52°, matrix 208 x 174, resolution 1.63×1.63mm, TR = 39.2ms, TE = 1.43ms, 25 phases per RR, averages 1.

### Coronary Point Source Evaluation

The use of larger k-space matrices in SNR-constrained acquisitions can theoretically result in little improvement in image quality if a large number of k-space coefficients are sampled when the signal is extremely low.^39^ To verify that the 0.85×0.85mm resolution images would result in meaningful increases in image quality, the led anterior descending (LAD) coronary artery in single-average b50 and b500 images in healthy subjects (n=7) was used as a point source. The location of the LAD was confirmed using bSSFP cine images (supplement, Figure S1). Diffusion-encoded images using the 64-channel coil were acquired at 0.85×0.85mm resolution and then immediately repeated at 2.5×2.5mm resolution. A region containing the led anterior descending (LAD) coronary artery was identified in the interventricular groove and rescaled up by 4x using bilinear interpolation (Matlab, Natick, MA).^40^ The perimeter (wall) of the coronary artery was segmented from the vessel lumen using a full-width-half max (FWHM) approach and the maximum diameter from each voxel on the perimeter was calculated.^40^ The median of these values was used as the final vessel diameter. The sharpness (mm^-1^) of the coronary wall was calculated as previously described,^40^ as the reciprocal of the shortest distance between voxels 20% and 80% above of the minimum intensity range. In each subject coronary diameter and sharpness were measured in a minimum of 3 single-average short axis slices and then averaged to yield a single per-subject value.

### SNR and Diffusion CNR calculations

SNR calculations with the 64 and 32 channel coils were performed using single-average b50 and b500 images in healthy volunteers (n=6). Noise images were acquired with the same sequence but with the transmit voltage set to zero, a repetition Jme of 2.6s and an average of 240 repetitions. The standard deviation of the signal in each voxel in the noise images was calculated to create a noise map. (In 1/6 subjects noise images were not available and the standard deviation of the voxels in a region-of-interest outside the body were used). SNR values were calculated by dividing the signal in each voxel by the standard deviation of the noise in that voxel and then averaged over all voxels in the septum. This was performed for each diffusion-encoding direction and on the full stack of short axis slices to derive a single average b50 and b500 SNR value for each individual. Diffusion contrast was defined as the difference in signal intensity in each voxel between the b50 and b500 images. The diffusion contrast-to-noise ratio (dCNR) was calculated in each voxel by dividing this difference by the standard deviation of the noise. An average dCNR value for each subject was calculated, as described above, across all diffusion encoding directions and short axis slices. SNR and dCNR maps with the 64 and 32-channel arrays are shown in the supplement (Figure S2). SNR and dCNR in the healthy volunteers were significantly higher with the 64-channel coil (supplement, Figure S2) and imaging of the AS and CTL subjects was, consequently, performed exclusively with the 64-channel array.

### Image Processing and Masking

Retrospective gating of the free-breathing diffusion-encoded images was performed using a low rank multitasking approach, as previously described.^12^ Derivation of the MD, FA and HA maps was performed with a cardiac-tailored in house version of Trackvis (Martinos Center, Boston MA) and further processed using Matlab. The endocardial and epicardial borders were determined using the b50, MD and HA maps and the generated masks were then applied to all maps derived from these images. Careful attention was paid to ensure that all endocardial and epicardial trabeculations were excluded from the boundaries. The center of the LV and the RV insertion points were manually identified, creating a polar coordinate system going radially from endocardium to epicardium and angularly (Θ) from the anterior to inferior RV insertion point. Voxels on a Cartesian axis were rebinned to a radial axis using bicubic interpolation to maintain the correct number of voxels per unit distance along each radial line. In many subjects the anterior and inferior walls were suitable for analysis, and in some cases most of the LV could be analyzed. However, data analysis in this study was limited to the voxels in the interventricular septum, where image quality was consistently high in all subjects. Diastolic LV thickness was measured in bSSFP cine images at the midventricular level that matched the central slice used in the DTI analysis. T1 maps were acquired in 3 short axis slices at the mid-ventricular level in each subject. An elliptical ROI was placed in the midseptum of each map and the T1 values in all 3 maps were averaged into a single per-subject value.

### HA Variance and Gradient Calculations

HA Variance was determined by separating the voxels within the septum of a single HA map into ten bins based on transmural depth. The bins were determined both by absolute depth in mm (Var_mm) and by %-transmurality (Var_%). The innermost and outermost bins were consistently noisy and they were, therefore, excluded from the analysis. The variance per bin was calculated for the remaining 8 bins and averaged into a single per-map variance value. The variance values in all HA maps analyzed in an individual subject were then combined to derive a single per subject variance value.

HA gradients were calculated for each voxel using its 8 neighboring voxels. The adjacent voxels on the radial line intersecting the target voxel were deemed radial, those perpendicular to the radial line circumferential, and the remaining voxels as oblique. For each neighboring voxel, the gradient was measured as the absolute change in HA from the target voxel divided by the distance between the voxels. Radial and circumferential HA gradients can be calculated bidirectionally (using the mean of the two radial/circumferential voxels) or unidirectionally (radially from endocardium to epicardium only, and circumferentially in a counterclockwise direction only). The two approaches yielded very similar results, but to avoid duplicate use of the same gradient in two adjacent target voxels, we chose to use the unidirectional approach. One limitation of this approach, however, is that no radial gradient can be calculated for the outermost (most epicardial) voxels in the heart. Minimum and maximum HA gradients were calculated by the smallest and greatest gradients between all 8 neighboring voxels, respectively. Mean gradient was defined as the average of all eight HA gradients and gradient range was determined by the difference between the maximum and minimum HA gradients. In addition, the relationship (radial, circumferential, or oblique) of the voxel producing the minimum and maximum HA gradients to the target voxel was recorded and then discretized into one of 3 possible orientation angles relative to the radial vector (radial = 0°, circumferential = 90°, oblique = 45°). The radial HA gradient was calculated as both an absolute gradient (degrees/mm) and a relative gradient (degrees/%-transmurality). All other gradients were calculated only as absolute gradients. The HA gradients were measured in all voxels within the septum in all the slices analyzed, and then averaged to yield a single per-subject value.

DTI-Tractography was performed in selected subjects using an in house cardiac-tailored version of Trackvis (Martinos Center, Boston MA). Cubic or spherical regions-of-interest (ROIs) were placed in the septum and cardiomyocyte orientation streamlines (tracts) intersecting the ROI were derived using a second-order Runge-Kutta algorithm. A tract termination angle of 35° was used and each segment (voxel) of the tract was color-coded by the HA in that segment.

### Principal Component and Cluster Analysis

DTI-phenomapping was performed using hierarchical clustering techniques that have been used to analyze genetic,^17, 18^ and clinical datasets,^19^ in subjects with cardiovascular disease. 10 metrics with voxel specific values were used: b50, MD, FA, radial gradient per mm, radial gradient per %-transmurality, circumferential gradient, mean gradient, minimum gradient, maximum gradient, and gradient range. Since the b50 images are highly T2-weighted, this was used as a surrogate for T2 within the voxel. The HA gradient values followed Gaussian distributions expect for the circumferential and minimum HA gradients. The absence of negative values and the low mean values of these parameters resulted in skewed distributions. To address this, all HA gradient values underwent square-root transformation, which resulted in highly Gaussian distributions suited to clustering (see supplement Figure S7). Each metric (column) in the voxel matrix then underwent z-score normalization. This can be done at the subject or population level, and the relative differences between these two approaches are extensively discussed in the supplement. As shown in the supplement (Figure S8) the differences between subject and population-based normalization were small. We chose to perform subject-based normalization, where a z-score is ascribed to each voxel based only on the voxel values in that subject, for the following reasons: The aim of subject-based normalization is to detect distinct microstructural microenvironments within individuals and to determine how reproducibly these microenvironments are found across the population. In addition, subject-based normalization resulted in more weighting of scalar based metrics, such as MD and FA, in the PCA and cluster analysis. While the results of PCA and cluster analysis on the subject and population-normalized data were highly similar (supplement Figure S8), the properties of subject-based normalization were better aligned with the aims of the phenomapping analysis in this study.

The subject-normalized voxels in the phenomapping matrix were analyzed using principal component analysis (PCA) and cluster analysis in Matlab. Ten principal components were identified, with the coefficients and contribution to total variance of each recorded. Cluster analysis was performed with k-means clustering, positioning centroids in a 10-dimensional space and minimizing total point to centroid distance. Clustering was performed using 1000 iterations and 20 replications to derive global minima. The number of clusters (k-value) chosen was based on the Davies–Bouldin index, which revealed similar minima at k=4 and k=5 in all matrices (clusters produced with k=5 are shown in supplemental Figure S8). However, for simplicity and initial demonstration of the approach, we focused principally on k=4. The properties of the voxels in each of the 4 clusters were assessed to determine which DTI-derived metrics most strongly characterized that cluster. Cluster analysis was performed on the entire population (AS + CTL) and in the AS and CTL subjects separately. To determine the impact of spatial resolution on the phenomapping approach the HA gradient maps were downsampled to a resolution of 2.5×2.5mm. Histograms of the downsampled HA gradient maps were substantially narrower than those of the original HA gradient maps (supplement, Figure S9). The narrower range of the downsampled histograms suggests that phenomapping would be less able to identify distinct clusters at 2.5mm vs. 0.85mm resolution.

### Statistical Analysis

All statistical analysis was done in Graphpad Prism Version 10, except for the correlation calculations, which was performed in Matlab. The normality of all datasets was confirmed using the Shapiro-Wilk and Kolmogorov-Smirnov tests in Prism. The relationships between all metrics available on a per subject level were quantified using Pearson correlation. Comparisons between two groups were performed using an unpaired, two-tailed t-test. If the data in the groups were related (e.g. high resolution vs. lower resolution versions of the same image/map) a paired, two-tailed, t-test was used. Comparisons between 3 or more groups were performed using ANOVA with Tukey post-test comparison. All data are reported and plotted as mean ± standard deviation. A p-value of < 0.05 was required to meet significance.

## Disclosures

Specific to this study: None

(General Disclosures: The Martinos Center for Biomedical Imaging has a research agreement with Siemens Medical. Dr. Elmariah is a consultant for Edwards Lifesciences, Medtronic, and Abbott and receives research funding from Edwards Lifesciences and Medtronic.)

## Data Availability

The data are available from the authors upon reasonable request.

## Funding

Funded in part by the following studies from the National Institutes of Health: R01HL159010, R01HL141563, R01HL151838

## Notes

### Competing Interest Statement

The authors have declared no competing interest.

### Funding Statement

This study was funded by the following grants from the National Institutes of Health (NIH): R01HL159010, R01HL141563, R01HL151838

### Author Declarations

The Institutional Review Board (IRB) of the Massachusetts General Hospital gave ethical approval for this work.

