## Supplement for "Diffusion Tensor Phenomapping of the Healthy and Pressure-Overloaded Human Heart"

**Supplement - Table of Contents**

1. MRI hardware and pulse sequence used for sub-mm DTI.
2. Diffusion contrast in the heart with 64 and 32-element radiofrequency arrays.
3. Voxel size and helix angle (HA) variance.
4. Comparison of cardiac DTI at 0.85mm and 2.5mm resolution.
5. Orientation of the minimum and maximum HA gradients.
6. Characteristics of aortic stenosis (AS) and age-matched control (CTL) subjects.
7. Conceptual basis of DTI-phenomapping.
8. Z-score normalization of DTI-derived data.
9. Subject vs. population-based normalization.
10. K=5 Clustering.
11. Impact of spatial resolution on DTI-phenomapping.

**Radiofrequency Coil and Pulse Sequence Used for Sub-mm Cardiac DTI.**

Free-breathing diffusion tensor MRI (DTI) of the heart with sub-mm resolution was performed using a diffusion-encoded spin-echo sequence with a spatially selective 2D excitation pulse and M2-compensated diffusion gradients, designed to null the first and second moments of motion (Figure S1).[1] A cardiac-tailored 64-channel array with uniformly arranged overlapping elements was used to acquire the data (Figure S1),[2] and retrospective respiratory gating of the diffusion-encoded images was performed with a low-rank multitasking approach.[1] The assessment of DTI image quality was performed using the cross section of the left anterior descending coronary artery (LAD) as a point source. A balanced steady state free precession (bSSFP) image of the heart and LAD at the same spatial resolution (0.85 x 0.85 x 8mm) provided a gold standard. As shown in Figure S1, the sharpness and clarity of the LAD were similar in the bSSFP, b50 and b500 images.

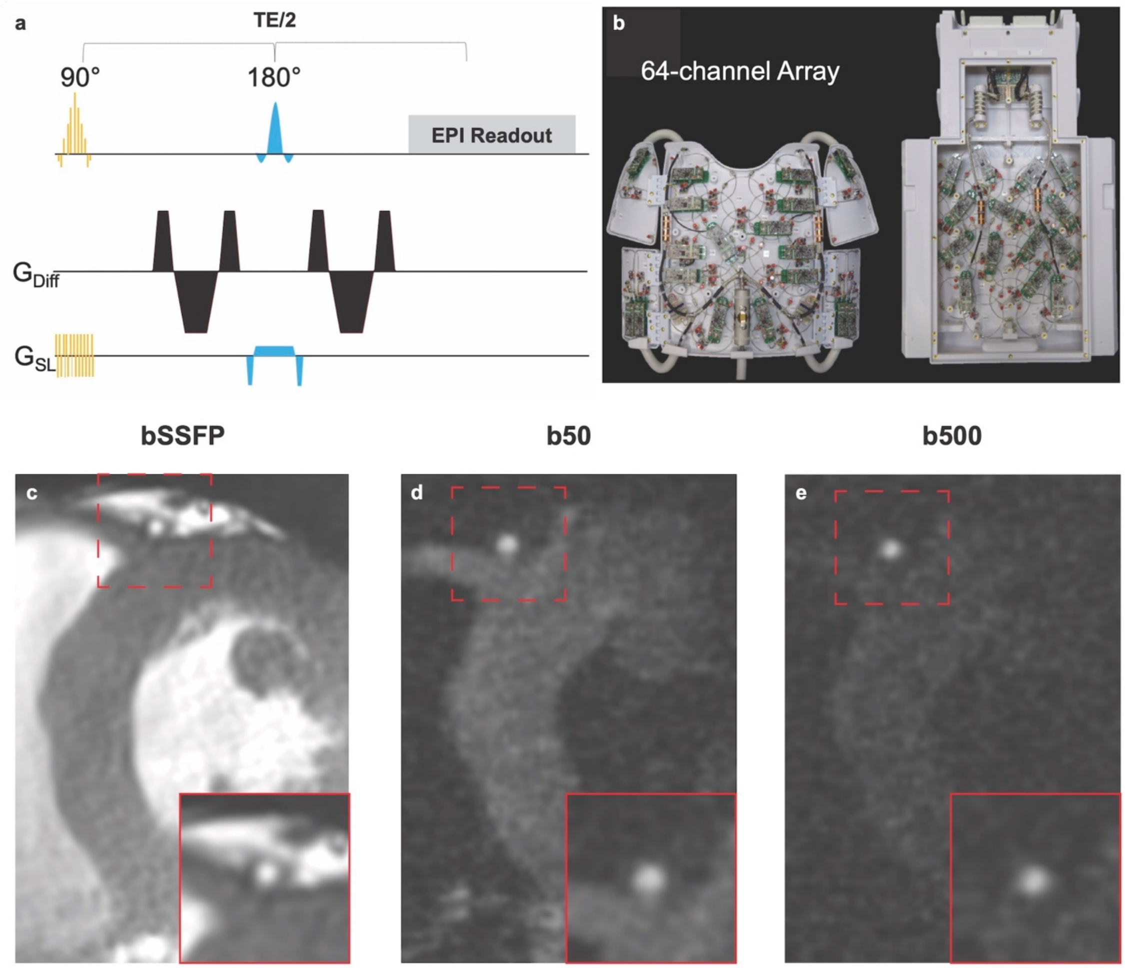


**Figure S1. Spatially selective excitation pulse and 64-channel radiofrequency coil used for sub-mm cardiac DTI.** (a) Diagram of spatially selective 2D excitation pulse and M2-nulled diffusion encoding gradients. The excitation pulse selectively excites an inner volume centered over the septum of the heart. M2-nulling prevents the motion of the heart, both at constant velocity (M1) and during periods of acceleration (M2), from corrupting the diffusion signal. (b) Anterior and posterior elements of the 64-channel cardiac array with the covers removed. The elements are 90mm in diameter and equally distributed in an overlapping geometry to minimize mutual inductance and the geometric or g-factor of the coil. (c-e) bSSFP, b50 and b500 images of the heart at the midventricular level showing the cross section of the LAD. All images were acquired with the 64-channel radiofrequency coil and a spatial resolution of 0.85 x 0.85 x 8mm. Magnified views of the area demarcated by the dashed red square are shown in the insets at the bottom-right of each image. The size, shape and clarity of the LAD are similar in all 3 images.

**64-Channel Radiofrequency Coil Facilitates Sub-mm Cardiac DTI.**

The details of the 64-channel radiofrequency coil have been previously described.[2] The physical dimensions of the coil allowed all the subjects in this study to be comfortably imaged. (The largest subject we have imaged with the coil to date was 1.9m tall and weighted 110 kg). In six of the healthy volunteers imaged during the technical development arm of the study detailed signal-to-noise maps comparing the 64-channel coil and a commercial 32-element array were obtained. Cardiac DTI was performed with the 64-element coil and, thereafter, the 32-element array at a resolution of 0.85 x 0.85 x 8mm. Noise images of both coils were acquired with the identical DTI sequence but with the transmit voltage set to zero, a repetition time of 2.6s, and an average of 240 repetitions.

The signal-to-noise ratio (SNR) in each voxel was calculated by dividing the signal in the b50 or b500 images by the standard deviation of the signal in that voxel on the noise images. Single average SNR maps were derived for each diffusion encoding direction and then averaged to obtain a final single-average SNR map for each slice. Final SNR maps acquired with the 64 and 32-element arrays are shown in Figure S2. Diffusion contrast to noise ratio (dCNR) maps were created by dividing the difference in signal between the b50 and b500 images in each voxel by the standard deviation of the noise in that voxel (Figure S2). The dCNR maps for each diffusion encoding direction were again averaged to obtain a final single-average dCNR map for each slice.

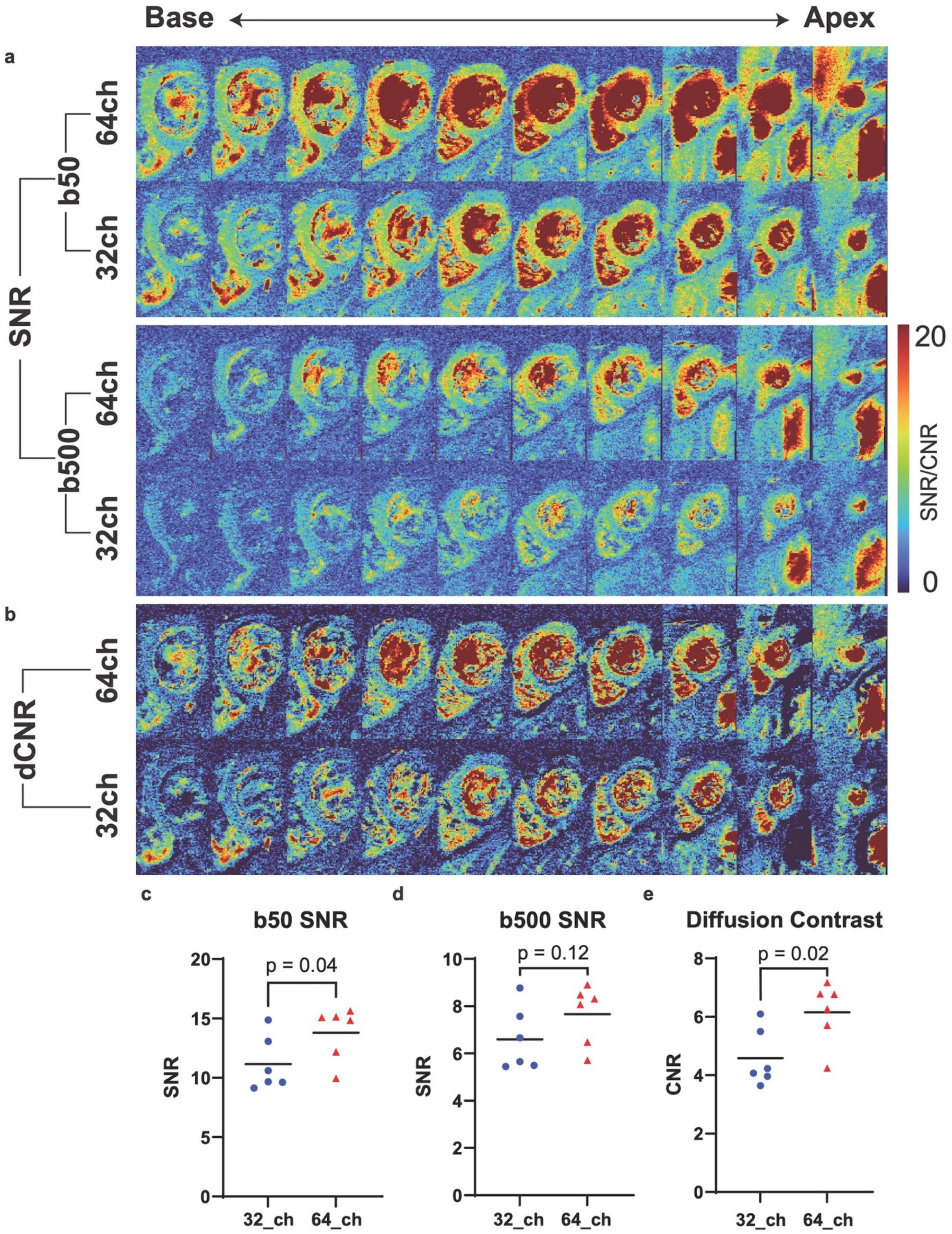


**Figure S2: Diffusion contrast in the heart is higher with the 64 vs. 32-element radiofrequency array.** (a) Single average signal-to-noise ratio (SNR) maps, at 0.85 x 0.85 x 8mm resolution, using the cardiac-tailored 64-channel coil and a commercial 32-channel array. SNR is highest in the septum and increases from base to apex. (b) Diffusion contrast-to-noise ratio (dCNR) maps, where diffusion contrast is defined by the difference in the signal between the b50 and b500 images. dCNR is highest is the septum at the midventricular level. (c) SNR in the septum with the 64-channel coil was significantly increased in the b50 images (14±2 vs. 11±2, p=0.04) and, (d) mildly increased in the b500 images (7.7±1.3 vs. 6.6±1.4, p=0.12). (e) dCNR was significantly increased with the 64-channel coil (6.2±1.1 vs. 4.6±1.0, p=0.02). For the comparisons shown in c-e, significance was determined with a paired t-test, n=6.

### Impact of Sub-mm Spatial Resolution.

#### Voxel Size and Helix Angle (HA) Variance

HA range from endo to epicardium increased mildly in the subjects with AS. In contrast, myocardial thickness and consequently the number of voxels from the endo to epicardium was markedly increased in AS. The HA range per voxel (or radial HA gradient) was, therefore, significantly lower in the AS subjects than the age matched controls (main text Figure 4). No significant differences were seen in HA variance between the AS and control (CTL) groups (main text Figure 3), however, a positive but non-significant correlation was seen between radial HA gradient and HA variance (main text Figure 5) in both groups. This raised the possibility of the HA variance values in AS being reduced by the lower radial HA gradient (increased number of transmural voxels) in these subjects, and not independently reflecting the degree of microstructural coherence in the myocardium. To investigate this effect, the HA maps in each AS subjects were downsampled so that the number of voxels from endo-to-epicardium matched the average number of transmural voxels in the CTL group. The downsampling factor used to create these ASmatch datasets averaged 1.42±0.13. No significant differences in HA variance were seen between the AS, ASmatch or CTL groups (main text Figure 3), confirming that the low HA variance values in AS accurately reflected the microstructural coherence of the myocardium in these subjects. In addition, as shown in Figure S3, no significant differences were seen between the AS and ASmatch datasets for any of the metrics used in this study.

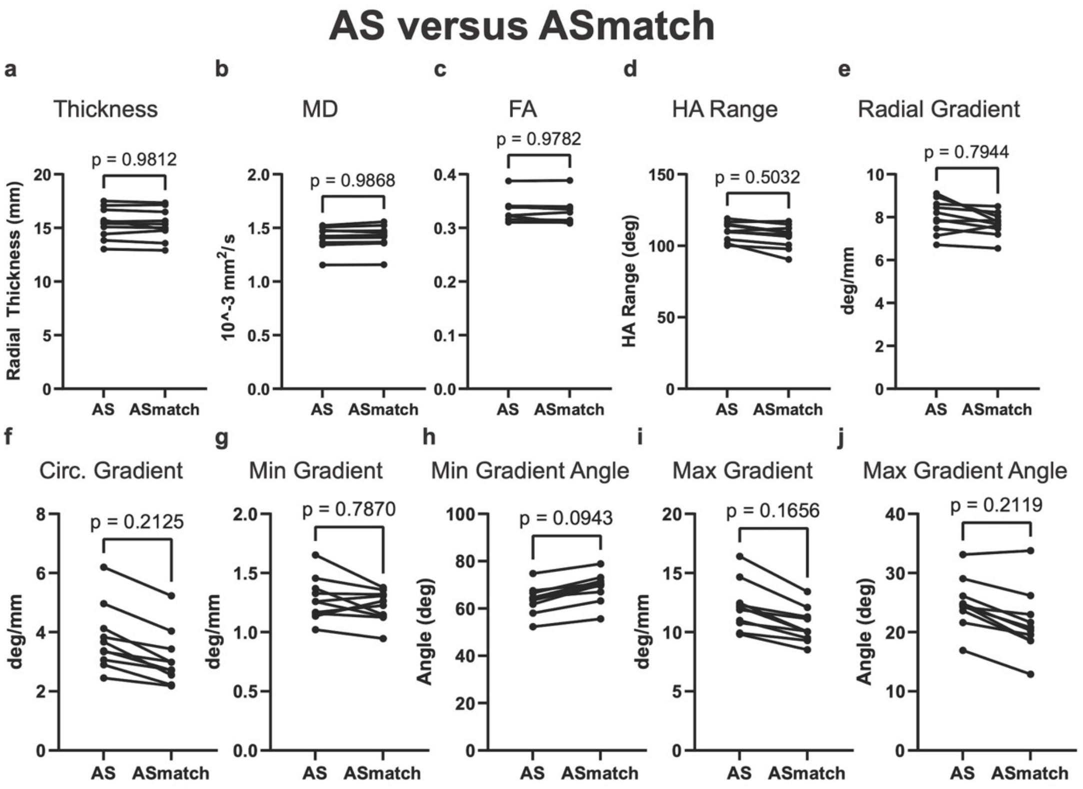


**Figure S3. DTI-derived metrics in the AS and ASmatch datasets.** Downsampling the voxels in the ASmatch datasets, so the number of transmural voxels equaled the average number of transmural voxels in the control subjects, did not significantly affect any of the metrics. Significance was determined by two-tailed paired t-test, n=10.

**Comparison of Cardiac DTI at Sub-mm and Standard Resolutions**

The DTI images in this study were acquired with an in-plane spatial resolution of 0.85 x 0.85mm, which results in a voxel almost 10-fold smaller than most current DTI studies (~2.5 x 2.5mm). To determine the impact of this improved resolution the MD, FA and HA maps in the CTL and AS groups were downsampled to have an in-plane resolution of 2.5 x 2.5mm. This represents a downsampling factor of 2.94, which is far higher than the average downsampling factor (1.4) used to create the ASmatch dataset.

Downsampling the images from 0.85mm to 2.5mm had a significant impact on the measurement of myocardial thickness and on all DTI derived metrics (Figure S4). In the CTL group significant differences between the original and downsampled images were seen in MD, FA, HA range, radial HA gradient, circumferential HA gradient, max gradient and the min/max gradient angles. In addition, a strong trend (p=0.07) towards difference was seen in the minimum HA gradient. Likewise, profound differences were seen in the AS subjects between 0.85mm vs. 2.5mm resolution (Figure S4). Significant differences were seen in MD, HA range, radial HA gradient, circumferential HA gradient, min gradient, max gradient, and the min/max gradient angles. In addition, a trend (p=0.1) towards difference was seen in FA. At the lower (2.5mm) resolution HA range and all HA gradients were reduced, consistent with a smoothing or averaging effect.

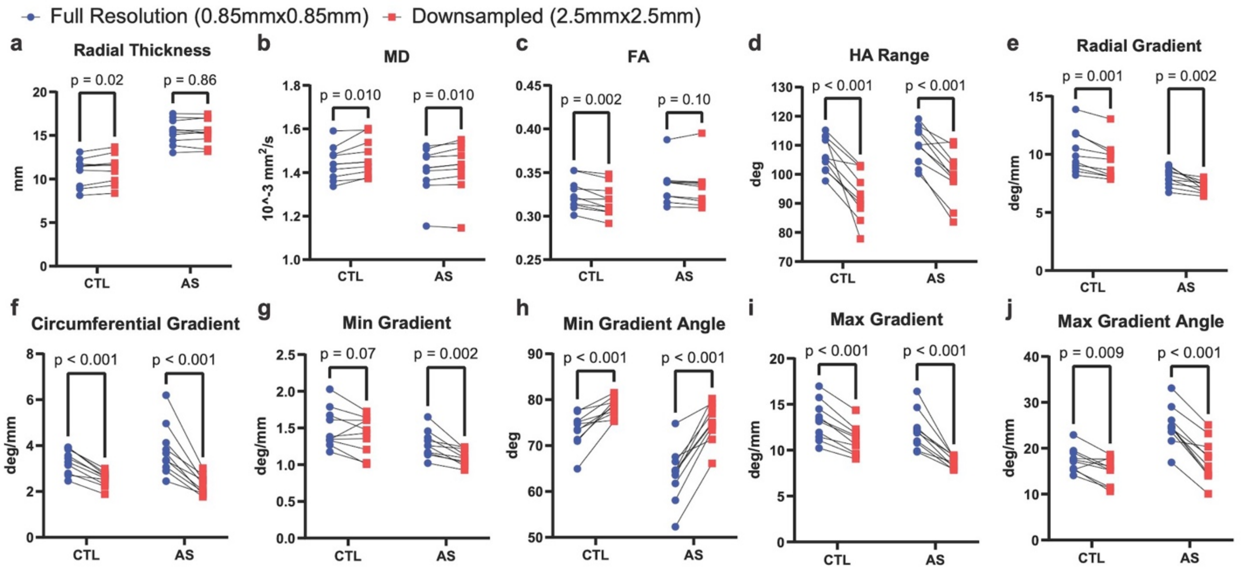


**Figure S4**. **Impact of downsampling images from full resolution (0.85mmx0.85mm) to standard resolution (2.5mmx2.5mm).** (a) The thickness of the myocardium differed significantly in the healthy controls at 0.85 vs. 2.5mm resolution (p=0.02). In the AS subjects, in whom the myocardium is substantially thicker, similar values were obtained at both resolutions. (b) MD was increased at the standard resolution in both the CTL (1.47±0.08 $10^{-3}mm^2/s$ vs. 1.44±0.08 $10^{-3}mm^2/s$, p=0.01) and AS (1.42±0.12 $10^{-3}mm^2/s$ vs. 1.41±0.11 $10^{-3}mm^2/s$, p=0.01) subjects. (c) FA was significantly reduced by downsampling in the CTL subjects (0.318±0.018 vs. 0.325±0.017, p=0.002) and trended lower in the AS subjects (0.334±0.024 vs. 0.336±0.021, p=0.1). (d) HA range was significantly reduced at 2.5mm resolution in both the CTL (93±9° vs. 107±6°, p<0.001) and AS (99±9° vs. 110±6°, p<0.001) subjects. (e) Likewise, radial HA gradient was significantly reduced at 2.5mm resolution in both the CTL (9.4±1.6°/mm vs. 10.2±1.8°/mm, p=0.001) and AS subjects (vs 7.2±0.5°/mm vs. 8.0±0.8°/mm, p=0.002). At the downsampled (2.5mm) resolution marked reductions in circumferential HA gradient were seen in the CTL (2.5±0.3°/mm vs. 3.3±0.5°/mm, p=0.001) and AS subjects (2.3±0.4°/mm vs. 3.8±1.1°/mm, p=0.002). (g) Minimum HA gradient trended towards a lower value in CTL (1.41±0.26°/mm vs. 1.50±0.27°/mm, p=0.07) and was significantly reduced in AS (1.10±0.11°/mm vs. 1.28±0.18°/mm, p=0.002). (h) Downsampling caused the minimum HA gradient to become more circumferential in both the CTL (78±2° vs. 73±4°, p<0.001) and AS subjects (75±4° vs. 63±6° p<0.001). (i) The maximum HA gradient was significantly reduced at standard resolution in the CTL (11.1±1.6°/mm vs. 13.2±2.1°/mm, p<0.001) and AS subjects (8.7±0.6°/mm vs. 12.1±2.1°/mm, p<0.001). (j) The angle of the maximum HA gradient became significantly more radially oriented at 2.5mm resolution in the CTL (15±3° vs. 17±3°, p=0.009) and AS subjects (17±5° vs. 25±4° , p<0.001). N=10 for all groups. All metrics are expressed as mean ± standard deviation per subject. P-values are the result of two-tailed paired t-tests.

These data underscore the importance of high spatial resolution in cardiac DTI, and complement the comparisons made between 0.85mm and 2.5mm resolution datasets in healthy volunteers in the development arm of the study (main text Figure 1). The importance of sub-mm resolution for the phenomapping approach we introduce is further demonstrated in Figure S8, below.

**Orientation of the Minimum and Maximum HA Gradients.**

DTI in the AS subjects did not show any evidence of a loss of microstructural coherence in the myocardium. However, the radial HA gradient (degrees/mm) was significantly reduced in AS, reflecting the mild increase in HA range but profound increase in LV thickness seen in these subjects (main text, Figure 4). In both the AS and CTL subjects the angle of the maximum gradient was most likely to be radial. Likewise, in both groups the angle of the minimum gradient was most likely to be circumferential. However, the proportion of oblique voxels accounting for the maximum and minimum gradients was higher in AS than CTL (main text, Figure 4). This is likely related to the lower radial HA gradient in AS, since as the radial gradient decreases and approaches the value of the circumferential gradient, oblique voxels become more likely to subtend maximum and minimum HA gradients (Figure S5).

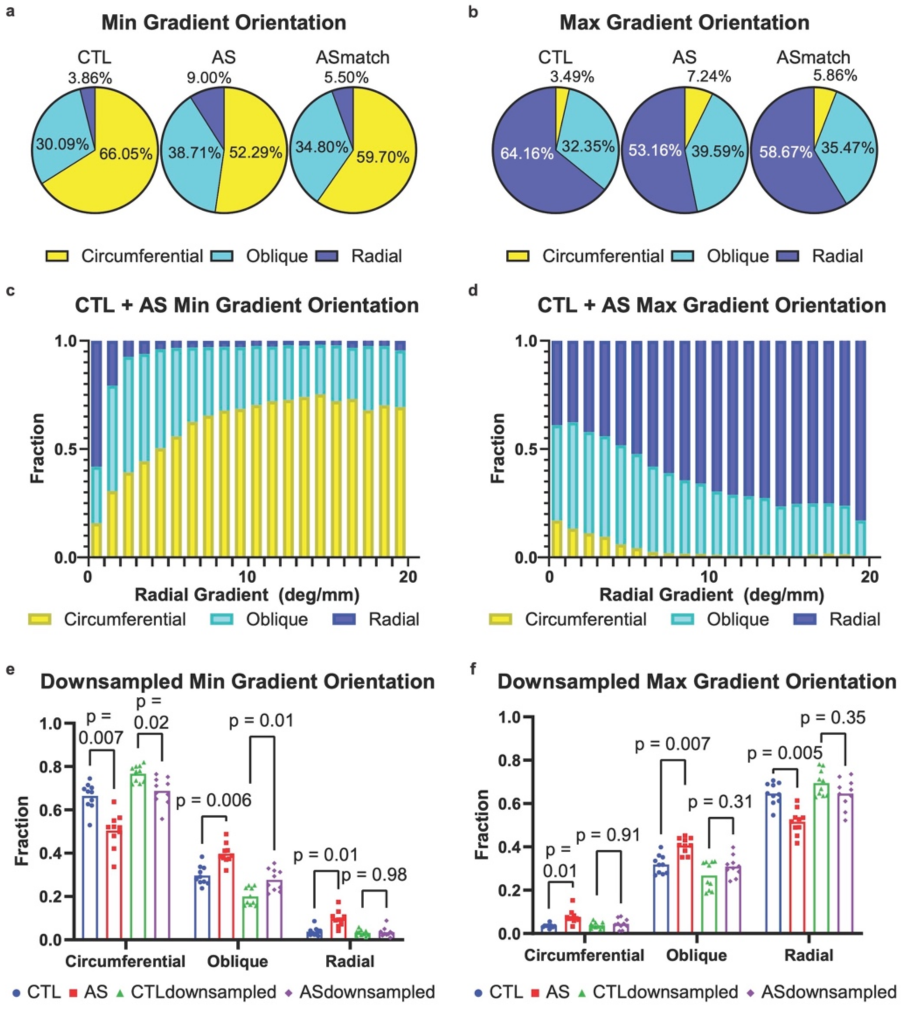

**Figure S5. Factors affecting the angle of the minimum and maximum HA gradients.** (a) The angle of the minimum HA gradient was less likely to be circumferential in the AS subjects vs. CTL. (b) Likewise, the angle of the maximum HA gradient was less likely to radial in the AS subjects. The distribution of these angles in the ASmatch dataset, however, was more similar to control. This raised the possibility of factors related to spatial resolution and/or radial HA gradient influencing the distribution of these angles. (c) The distribution of minimum gradient angle as a function of radial HA gradient is shown. As the radial HA gradient decreases, the minimum gradient angle becomes less likely to be circumferential and more likely to be oblique. (d) Likewise, as the radial HA gradient decreases, the maximum gradient angle becomes less likely to be radial and more likely to be oblique. (e, f) Distribution of min and max gradient angles in HA maps with original (0.85mm) resolution and those downsampled to 2.5mm resolution. At 2.5mm resolution the differences in the distribution of the min HA gradient angle between AS and CTL are reduced but remain significant. In contrast, differences in the max HA gradient distribution between AS and CTL lose all significance at 2.5mm resolution.

### Characteristics of AS and CTL Subjects.

The AS and CTL subjects were well matched and no significant differences were seen in age, sex, body mass index (BMI) and heart failure between the two groups (Table S1). A trend towards increased hypertension was seen in the AS subjects but did not reach significance. Hypertension, when present, was well controlled and below target levels in all but 1 case. Diabetes was present in 50% of the subjects with AS but was well controlled with a mean HbA1c of 6.05. (HbA1c data were not available in all the CTL subjects and are, therefore not reported).

In the AS subjects 80% had tricuspid aortic valves and 20% bicuspid valves (Table S1). The average values for the peak and mean aortic gradients were 76 mmHg and 44 mmHg, respectively. The highest peak gradient was 94mmHg and no AS subjects had a gradient > 100mmHg. The mean aortic valve area was 0.71 $cm^2$ and mean LV ejection fraction (EF) was 68%. The lowest EF value was 57% and 8/10 subjects had EF values >60%. The mean septal thickness by echo was 13.4mm. These parameters are consistent with severe but well compensated AS and, in the absence of symptoms, aortic valve replacement would not be indicated according to current guidelines.[3, 4]

**Table S1.** Characteristics of Aortic Stenosis (AS) and Age-Matched Control (CTL) Subjects

| **Characteristic** | **AS (N = 10)** | **CTL (N = 10)** | **p-value** |
|---|---|---|---|
| Age – years (mean ± SD) | 78 ± 8 | 74 ± 8 | 0.3444 |
| Female sex – No. (%) | 6 (60%) | 7 (70%) | 0.7815 |
| BMI | | | |
| Median | 27.5 | 23.4 | 0.1230 |
| Interquartile range | 25.6-28.7 | 22.0-26.3 | |
| Significant CAD – No. (%) | 2 (20%) | 0 (0%) | 0.4737 |
| Prior MI – No. (%) | 1 (10%) | 0 (0%) | 0.1000 |
| GFR (ml/min) – Mean ± SD | 61.9 ± 16.7 | -- | |
| Heart Failure (HFpEF) – No. (%) | 0 (0%) | 0 (0%) | 1.0000 |
| Heart Failure (HFrEF) – No. (%) | 0 (0%) | 0 (0%) | 1.0000 |
| Hypertension | 9 (90%) | 4 (40%) | 0.0573 |
| Well Controlled – No. (%) | 8 (80%) | 4 (40%) | |
| Not controlled – No. (%) | 1 (10%) | 0 (0%) | |
| Hypertension Treatment | | | |
| ACE Inhibitor – No. (%) | 2 (20%) | 3 (30%) | 1.0000 |
| ARB – No. (%) | 4 (40%) | 1 (10%) | 0.5820 |
| Diuretic – No. (%) | 4 (40%) | 1 (10%) | 0.3034 |
| Calcium Channel Blocker – No. (%) | 4 (40%) | 1 (10%) | 0.3034 |
| Beta Blocker – No. (%) | 4 (40%) | 0 (0%) | 0.0867 |
| Diabetes – No. (%) | 5 (50%) | | |
| HbA1c (%) – Mean ± SD | 6.05 ± 0.81 | -- | |
| Diabetes Treatment | | | |
| Metformin – No. (%) | 2 (20%) | -- | |
| Insulin – No. (%) | 1 (10%) | -- | |
| SGLT2 – No. (%) | 1 (10%) | -- | |
| None – No. (%) | 1 (10%) | -- | |
| *Echo Characteristics* | | | |
| Valve Morphology | | | |
| Bicuspid – No. (%) | 2 (20%) | -- | |
| Tricuspid – No. (%) | 8 (80%) | -- | |
| Peak Gradient (mmHg) – Mean ± SD | 76.40 ± 12.96 | -- | |
| Mean Gradient (mmHg) – Mean ± SD | 43.60 ± 6.92 | -- | |
| Aortic Valve Area ($cm^2$) – Mean ± SD | 0.71 ± 0.14 | -- | |
| Ejection Fraction (%) – Mean ± SD | 67.5 ± 7.3 | -- | |
| LVH – No. (%) | 7 (70%) | -- | |
| Interventricular Septum Thickness (mm) – Mean ± SD | 13.4 ± 2.1 | -- | |

**Abbreviations used in Table S1.** *BMI = body mass index, CAD = coronary artery disease, MI = myocardial infarction, GFR = glomerular filtration rate, HFpEF = heart failure with preserved ejection fraction, HFrEF = heart failure with reduced ejection fraction, ACE = angiotensin converting enzyme, ARB = angiotensin receptor blocker, HbA1c = hemoglobin A1c, SGLT2 = sodium-glucose cotransporter-2 inhibitor, LVH = left ventricular hypertrophy.*

### Diffusion Tensor Phenomapping.

Cardiac DTI data have hitherto been analyzed on a per-subject or per-segment basis, as shown in Figure 5 of the main text. Here we introduce a new formulism for the characterization of myocardial microstructure based on the determination of voxel-wise associations. The approach is analogous to single cell RNA-sequencing in a tissue specimen,[5, 6] where principal component analysis and hierarchical clustering of gene expression allows a variety of cells and cellular microenvironments to be defined.[5, 6] Here we consider each voxel much like an individual cell/nucleus and the array of DTI-derived metrics analogous to an array of gene expression (Figure S6). Principal component analysis and hierarchical clustering of the voxel matrix allows consistent associations to be identified and a range of microstructural micro-environments to be defined.

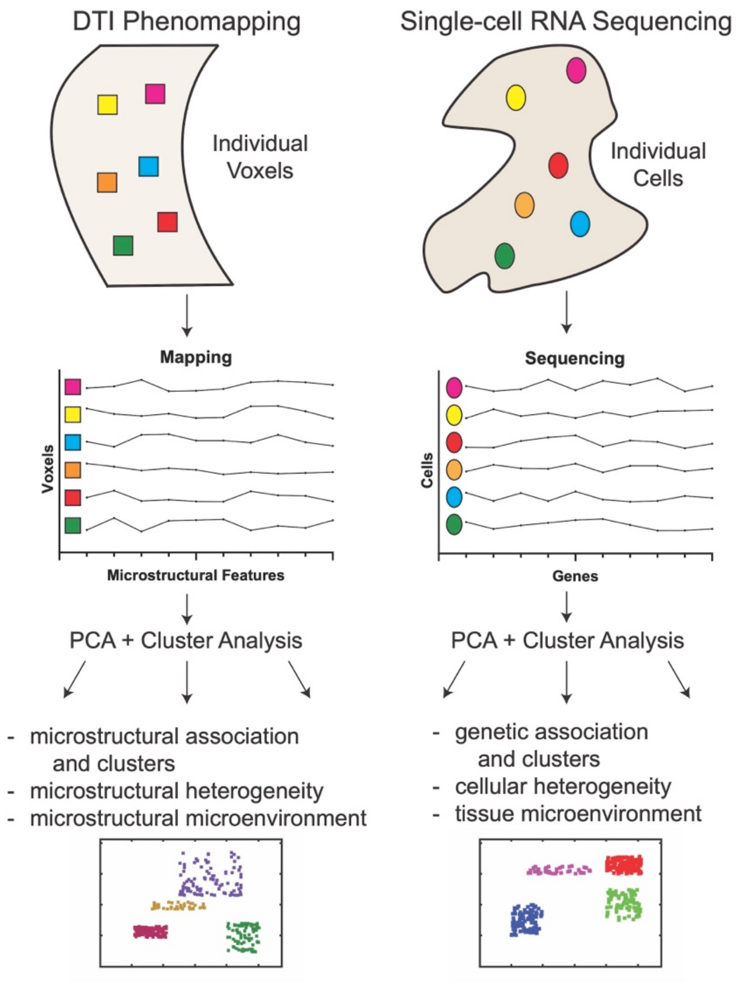


**Figure S6. Conceptual basis of DTI-Phenomapping.** The approach is similar to single cell RNA-sequencing of a tissue sample, where the population of cells is analogous to a population of voxels. In each voxel an array of DTI-derived characteristics is obtained, analogous to an array of gene expression in cells. The voxel and gene expression matrices are both suited to principal component analysis and hierarchical clustering. RNA-sequencing and DTI-phenomapping both allow local clusters and micro-environments with distinct properties to be identified. With RNA-sequencing distinct cellular micro-environments in a tissue can be detected. Similarly, DTI-phenomapping allows distinct microstructural environments to be detected.

### Normalization for DTI Phenomapping

The phenomapping approach requires all parameters in the matrix, which may have very different magnitudes and ranges, to be mapped to a common scale and to be given equivalent weight. A common approach used to do this involves z-score normalization, which assigns each value of a specific parameter a new value based on its standard deviation from the mean.[7, 8] Z-score normalization works optimally on parameters with a Gaussian distribution and is not recommended for skewed distributions. As shown in Figure S7, many of the DTI-derived parameters followed Gaussian distributions, however, some of the gradient-related parameters did not. This was most marked with the minimum and circumferential HA gradients, which both have very low mean values and no negative values, producing a skewed distribution. To address this, all gradient related values were square-root transformed prior to normalization, which resulted in highly Gaussian distributions (Figure S7) that were well suited to z-score normalization.

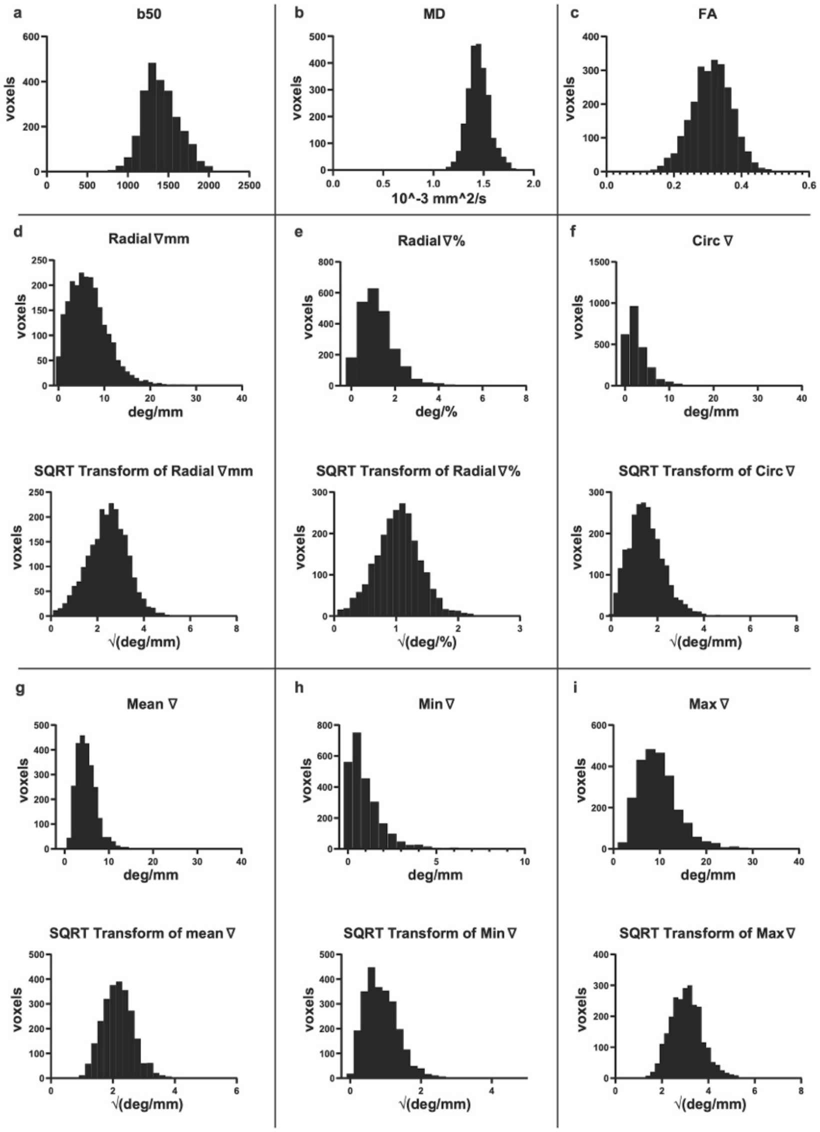


**Figure S7**. **Gaussian distribution of DTI-derived metrics facilitates Z-score normalization.** All histograms shown are of the full population (AS + CTL). (a-c) Scalar indices such as (a) b50 signal intensity, (b) MD and (c) FA have highly Gaussian distributions. (d-i) Some of the gradient-derived metrics, however, (e.g. circ and min gradients) showed skewed distributions. Taking the square-root of these HA gradient values transformed them from skewed to Gaussian distributions. The scalar (a-c) and square-root transformed gradient metrics (d-i) are all highly suited to Z-score normalization.

Z-score normalization can be performed on a per subject basis or at the level of the entire population (all subjects). Normalization at each of these steps addresses different questions. We elected here to perform normalization on a per subject basis (i.e. in all voxels from all slices in the same individual). Our goal in doing this was to determine whether certain microstructural associations, defining local microenvironments, would be consistently seen within an individual. We further aimed to determine whether these local microstructural patterns/environments would be consistently seen in all subjects in the population, including healthy subjects. Normalization at the population level (i.e. all voxels from all subjects) probes whether local deviations in one parameter across the entire population are associated with similarly large deviations in other microstructural parameters across the population.

**Individual vs. Population Based Normalization**

To further investigate the impact of normalization we performed principal component analysis and hierarchical clustering on the voxel matrix using either subject or population-based normalization. The b50 images, used in the subject-normalized phenomapping matrix, are highly T2-weighted and provide a useful measure of T2. The intensity of the b50 images, however, can differ markedly between individuals in the population due to differences in body habitus and coil loading. Population based normalization, therefore, required the b50 images to be removed to avoid large differences in signal intensity biasing the analysis. This is not an issue when subject-based normalization is performed since the b50 images in each subject are matched to a common scale. For completeness, however, principal component (PC) and cluster analysis with population-based normalization was compared to subject-based normalization both with and without the b50 data included (Figure S8). In all three cases the first 3 principal components accounted for >75% of total variation within the matrix and 4 distinct clusters could be identified.

Inclusion or exclusion of the b50 data did not change any of the principal components in the subject-normalized matrices (Figure S8a-b). In both cases PC1 was dominated by high radial, mean and max HA gradients; PC2 by high MD and low FA; and PC3 by high circ and min HA gradients. No major differences were seen between the principal components of the matrices with subject-based or population-based normalization (Figure S8c). Significant differences, however, were seen in the cluster analysis based on the normalization approach. Elimination of the b50 data did not significantly change the nature of the 1st, 2nd or 4th clusters in the subject-normalized matrices (Figure S8d-e). However, in the 3$^{rd}$ cluster elimination of the b50 images reduced the relative weighting of the other scalar values (MD and FA) and increased the weighting of the circumferential and minimum HA gradients (Figure S8d-e). The overall nature of the clusters did not differ significantly between the subject and population-normalized matrices, from which the b50 images had been removed (Figure S8e-f), however, the relative frequency of the various clusters did change. In the population-normalized matrix the voxels in the most abundant cluster (cluster 1) were now highly ordered and resembled the voxels constituting cluster 2 in the subject-normalized matrix. Likewise, the properties of the voxels in cluster 2 of the population-normalized matrix were extremely similar to those in cluster 1 of the subject-normalized matrix. No significant differences were seen in clusters 3 and 4 between the subject and population-normalized matrices (Figure S8e-f).

The percentage of voxels in each cluster with the various normalization approaches is shown in Figure S8g-i. With subject-based normalization, which weights differences within individual subjects and attenuates large differences (outliers) across the population, the ranges in each cluster were fairly narrow (Figure S8h). With population-based normalization, which strongly weights large differences across the population, the percentage of voxels in each cluster had a far broader range (Figure S8i). Overall, no major differences were seen in the percentage composition of the clusters between the AS and CTL subjects regardless of the normalization method used.

Collectively, these data suggest that subject vs. population-based normalization is unlikely to result in large differences, and that the two approaches are complementary. With both approaches the first 3 principal components accounted for >75% of total variation and were extremely similar. In addition, differences in the percentage of voxels in the various clusters were small and their properties mirrored each other extremely closely. Eliminating the b50 images from the subject and population-normalized matrices, however, did reduce the weight of the remaining scalar metrics (MD and FA) in the cluster analysis so that cluster 3 was no longer dominated by MD and FA. A full exploration of the characteristics of subject vs. population-based normalization, and the weighting of scalar vs. gradient based metrics, is beyond the scope of this paper and will require numerous additional studies. Our choice of subject-based normalization in this study (main paper Figure 6) was based on the following: 1) Ability to detect local microstructural microenvironments within individuals, including healthy subjects, 2) balanced weighting of scalar and gradient-based metrics in the cluster analysis so that cluster 3 was in large part determined by MD and FA and 3), narrower ranges of cluster composition in the AS and CTL subjects with subject-normalization. However, with either approach no major differences were detected between the AS and CTL subjects, consistent with other analysis methods used in the paper.

**K=5 Clustering.**

The number of clusters selected was determined using the Davies-Bouldin index, which showed similar minima at k=4 and k=5 in all cases. For simplicity and initial demonstration of the approach, we focused on k=4. However, clusters produced with k=5 are shown in Figure S8. Clustering was performed, as before, on the full voxel matrix (CTL + AS, Figure S8j) and then separately using those only voxels from the CTL (Figure S8k) or AS (Figure S8l) subjects. The cluster sets produced in all three cases (CTL + AS, CTL, AS) were extremely similar.

The pattern of the clusters produced with k=4 and k=5 showed a high degree of overlap. In both conditions clusters of highly ordered, least ordered and mixed voxels were observed. Moreover, the properties of these corresponding clusters had extremely similar characteristics at k=4 and k=5. However, the ordered cluster at k=4 (Figure S8d) was split into two clusters with k=5 (Figure S8j-l). One of these ordered clusters was characterized by average radial + max HA gradients and above average circ + min HA gradients. The other ordered cluster was characterized by above average radial + max HA gradients and below average circ + min HA gradients. The net effect of k=5 clustering was, therefore, to increase the impact of the circ and min HA gradients and split the ordered cluster into two. Further study will be required to determine the optimal number of clusters for DTI-phenomapping of the human heart, which could depend on the specific context and disease being studied.

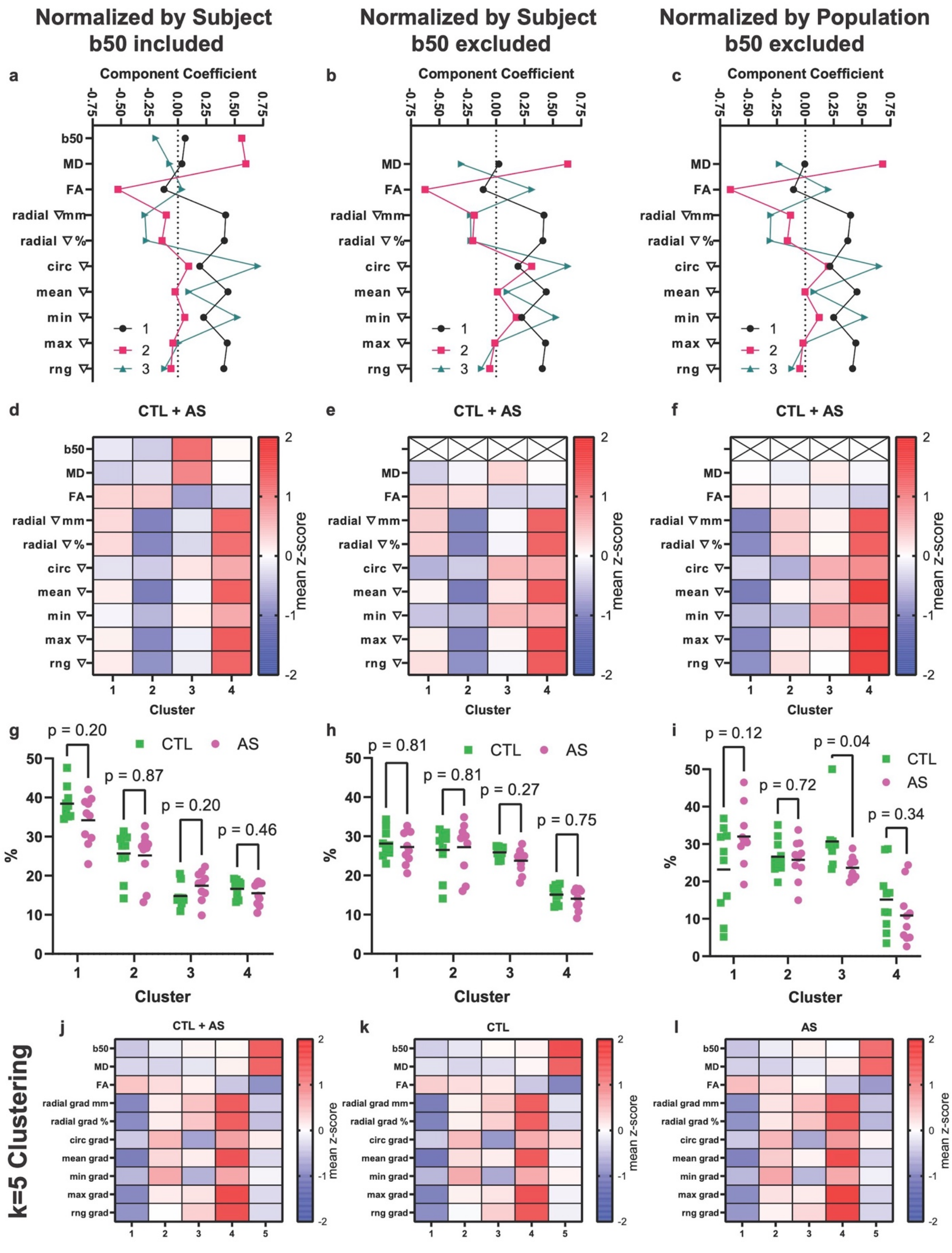


**Figure S8. Subject vs. population-based normalization for DTI-phenomapping.** (a) Principal component analysis (PCA) of the population matrix using subject-based normalization, as shown in the main manuscript (Figure 6). (b) PCA using subject-based normalization but with the b50 images excluded and, (c) PCA of the matrix using population-based normalization, also with the b50 images excluded. (a-c) In all cases the first 3 principal components (PCs) accounted for > 75% of variation and were extremely similar, regardless of the normalization approach used. (d) Hierarchical clustering using subject-based normalization, as shown in the main manuscript (Figure 6). (e) Exclusion of b50 data reduced the weighting of the remaining scalar metrics (MD, FA) and increased the weighting of the HA gradient metrics, particularly the circ and min HA gradients. This did not significantly change the composition of the 1$^{st}$, 2$^{nd}$ or 4$^{th}$ clusters. (e) However, the 3rd cluster was no longer driven by MD and FA but rather the circ and min HA gradients. (f) Population-based normalization did not significantly affect the characteristics of the clusters, but did alter their relative frequencies. With population-based normalization, highly ordered voxels with low HA gradients became slightly more frequent than voxels with average degrees of order. (g-i) The percentage contribution of each cluster in individual AS and CTL subjects is shown. No major differences based on the normalization approach were seen. However, with population-based normalization (i), the percent-contribution of the most ordered and least ordered clusters (1 and 4) became far more variable. Statistical significance was determined by two-tailed unpaired t-tests, n=10 per group. (j-l) Clustering with k=5 in the full voxel matrix (CTL + AS), CTL matrix and AS matrix, respectively. The cluster sets produced in all 3 cases were extremely similar.

**DTI Phenomapping is Optimally Performed at Sub-mm Resolution.**

The spatial resolution of the cardiac DTI images has a significant impact on the tensor-derived metrics, as shown in Figure S4 above. The metrics in Figure S4 are reported on a per-subject basis, where the individual values in all voxels are averaged to create a single value for each subject. DTI-phenomapping, however, is performed on a per voxel basis. We, therefore, aimed to determine whether the spatial resolution of the DTI images would have a similar effect on the phenomapping approach. We hypothesized that images acquired at the standard 2.5mm resolution would produce a narrower range of HA gradients and make it more difficult to distinguish separate clusters. To determine this, histograms of radial, circumferential, min and max HA gradients were derived using the original HA maps (0.85mm resolution) and HA maps downsampled to 2.5mm resolution. The histograms were square-root transformed to create Gaussian distributions based on the standard protocol used in this study. Representative histograms of a single subject and of the entire population (10 AS + 10 CTL ) are shown in Figure S9. In all cases, downsampling to 2.5mm resolution narrowed the variance of the HA gradient values. This effect was particularly marked for the radial, circumferential and maximum HA gradients. The results of this analysis confirmed our hypothesis and demonstrate that DTI phenomapping at sub-mm resolution results in a greater variance of HA gradient values and is more likely to facilitate the identification of distinct microenvironments and clusters. This adds further rationale for the ultra-high resolution approach used in this study.

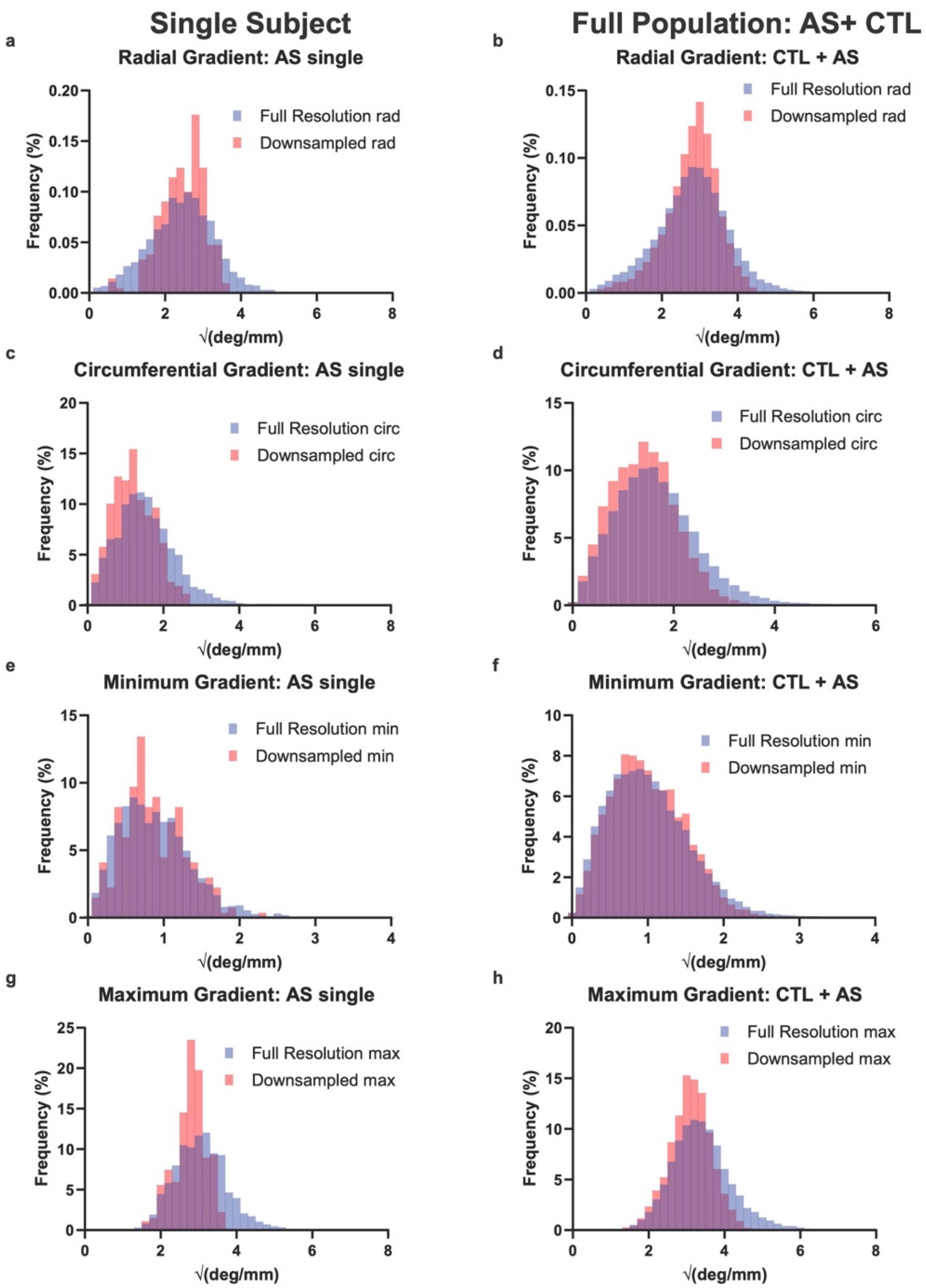


**Figure S9. Impact of spatial resolution on DTI-phenomapping.** Histograms of square-root transformed HA gradient values derived from HA maps at full (0.85x0.85mm) and downsampled (2.5x2.5mm) resolution are shown. Full resolution histograms are in blue, downsampled histograms are in red and areas of overlap are in purple. (a-h) Histograms from a single subject are shown on the left and from the entire population on the right. (a, b) The dynamic range of radial HA gradient values was markedly narrowed by downsampling to 2.5mm. Likewise, (c, d) the dynamic range of circumferential HA gradient values was reduced by downsampling but, (e, f) minimum HA gradient was less affected. (g, h) Downsampling markedly reduced the dynamic range of maximum HA gradient values.
